# Genetic Associations with Temporal Modeling of Alzheimer’s Disease Progression Supports a Novel Paradigm for Disease Risk

**DOI:** 10.64898/2026.07.07.26356710

**Authors:** Daniel M. Jordan, Eli Kritzer, Ryan C. Thompson, Anina N. Lund, Tulsi Patel, Ariela Buxbaum Grice, Divya A. John, Alzheimer’s Disease Neuroimaging Initiative, Alzheimer’s Disease Metabolomics Consortium, Alzheimer’s Disease Sequencing Project, Alison M. Goate, Benjamin S. Glicksberg, Negar Golestani, Maria Koromina, Alan E. Renton, Noam D. Beckmann

## Abstract

A major challenge in Alzheimer’s disease (AD) research is predicting who will develop AD, how it progresses, and how to slow, prevent, or reverse progression. Here, we apply a data-driven timeline inference framework to sparse longitudinal blood metabolomics data to reconstruct AD timelines and derive individual-specific timeline progression rates. Inferred temporal locations for each metabolomics sample along the AD timeline closely track clinical severity, while timeline progression rates capture inter-individual differences in the speed of pathophysiological progression. Genome-wide association studies of timeline progression rate identify novel loci distinct from those in AD case-control studies, notably showing no effect of the major risk locus *APOE*. These findings support a multidimensional paradigm of AD risk in which disease potential and progression act as partially independent factors. By explicitly modeling disease dynamics, this work reveals genetic contributions not captured by traditional approaches and provides a framework for studying AD and other progressive disorders.

## Introduction

Alzheimer’s disease (AD) is the most prevalent neurodegenerative disorder, currently affecting over 7 million Americans, a figure projected to double by 2060 without the development and widespread availability of effective disease-modifying therapies^1^. AD is a progressive disease characterized by a long preclinical phase followed by cognitive decline^2,3^. Years before clinical symptoms such as mild cognitive impairment (MCI) emerge, a stereotyped sequence of pathological changes unfolds, measurable through neuroimaging and fluid biomarkers^4–10^. The consistent ordering of events in this sequence suggests that AD develops along a definable biological timeline of disease.

AD is highly heritable^11,12^. Numerous loci associated with observed case-control status have been identified across multiple populations^13–17^. Among these, *APOE* is the strongest genetic factor influencing both case-control status and age at onset^18–21^. However, despite AD being an inherently progressive disorder, most large genetic studies of AD are cross-sectional. As a result, they have focused on observed AD case-control status rather than progression. Although this approach has been instrumental in identifying genetic drivers of disease susceptibility, it offers limited insight into the genetic factors that influence variation in clinical disease course among affected individuals. This limitation is particularly important because individuals with MCI and AD exhibit substantial heterogeneity in rate of clinical decline^22–24^, with some declining rapidly while others progress more slowly, and because predicting progression is critical for anticipating loss of independence and future care needs^25^. Rapidly progressing AD (rpAD) has been described as a distinct subtype^26,27^, and increasing evidence supports rpAD as one extreme in a continuum of progression rates^28,29^. AD progression rate has been suggested to be heritable^30,31^. However, several previous efforts have been unable to identify robustly associated loci, indicating the genetic basis of disease progression remains poorly understood and larger sample sizes or alternative study designs are needed^30–33^. Moreover, different patterns of progression do not correlate with *APOE* genotype^21^. Together, these suggest that the genetic factors influencing progression are at least partially distinct from those underlying the potential of having the disease, thereby motivating a thorough investigation of the genetic basis of AD progression.

To better understand the genetic architecture of AD progression, we use a data-driven timeline inference approach that we previously developed^34^. This framework leverages longitudinal high-dimensional molecular data to estimate each individual’s temporal location along the AD timeline and to define a quantitative trait for progression rate for each individual. Inferred AD timelines are not trained on any clinical measure or established biomarker, and thereby do not capture any single component of disease pathophysiology such as amyloidopathy, tauopathy or cognitive decline, but rather represent a more general continuum of disease progression across all disease aspects. Critically, this approach incorporates an intrinsic performance metric that enables evaluation of the inferred timeline without requiring a predefined reference event or known biomarkers. Our inferred temporal locations align with expected progression of clinical severity, while inferred timeline progression rate captures the rate of progression of these same measures of severity. This enables us to conduct a genome-wide association study (GWAS) to characterize the genetic basis of AD progression rate, rather than of observed case-control status as in traditional AD genetic studies. Using this approach, we discover new loci potentially implicated in AD progression rate, which are partially distinct from known AD risk loci, most notably being completely independent of the most prominent AD risk locus *APOE*. More broadly, this conceptual framework (**Figure 1**) provides a foundation for understanding biological drivers of disease heterogeneity, opens new strategies for modeling progressive and degenerative disease, and supports a multidimensional paradigm for the risk of AD that distinguishes genetic risk for AD potential from genetic risk for rate of AD progression.

**Figure 1.**
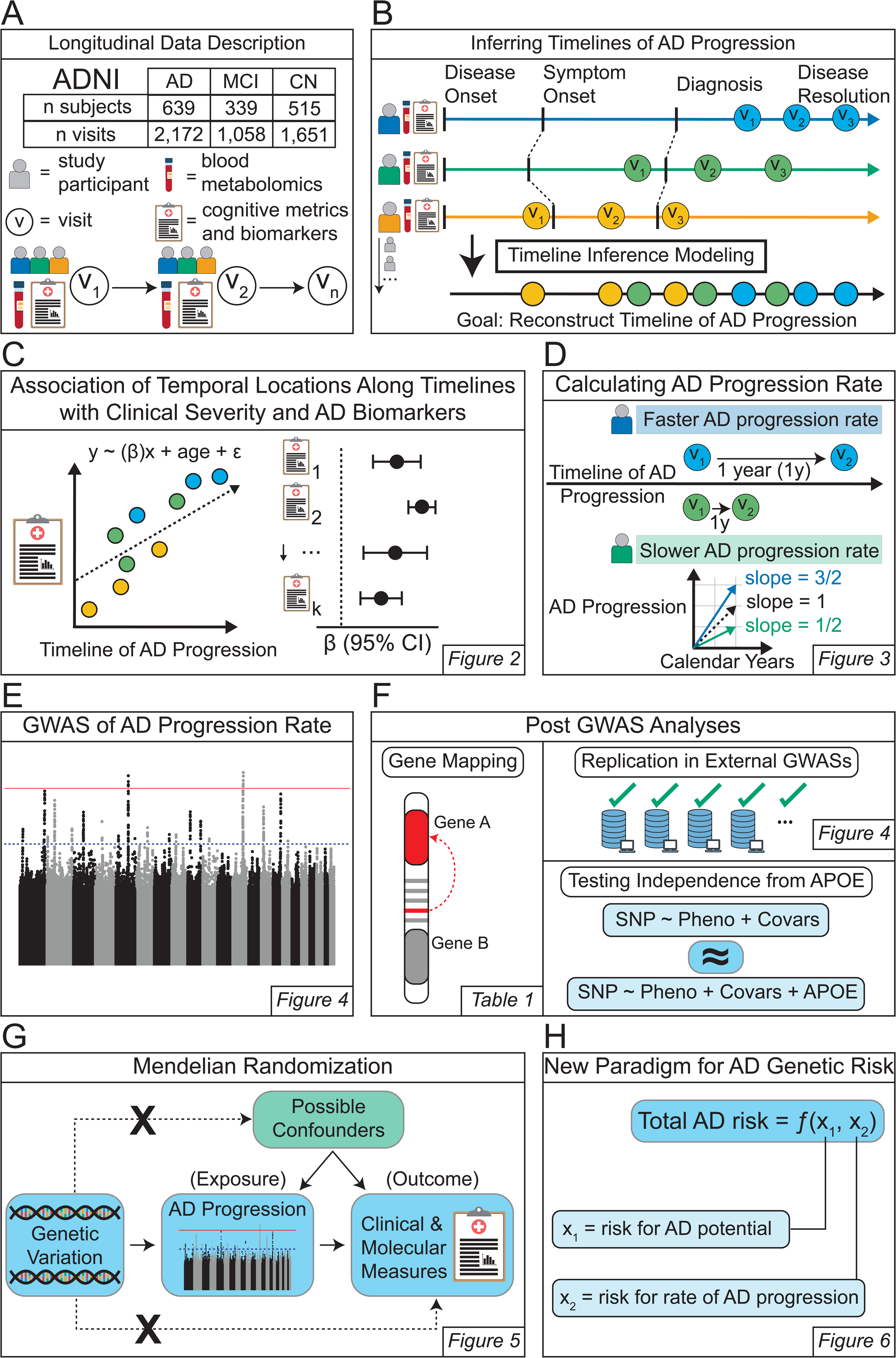
Schematic Overview. **(A)** Longitudinal Data Description. **(B)** Inferring Timelines of AD Progression. **(C)** Association of Temporal Locations Along Timelines with Clinical Severity and AD Biomarkers. **(D)** Calculating AD Progression Rate. **(E)** Genome-Wide Association Study (GWAS) of AD Progression Rate. **(F)** Post GWAS Analyses (Gene Mapping, Replication in External GWASs and Testing Independence From APOE). **(G)** Mendelian Randomization. **(H)** New Paradigm for AD Genetic Risk.

## Results

### The Alzheimer’s Disease Neuroimaging Initiative (ADNI) dataset

This study uses data from the ADNI dataset, a longitudinal cohort including participants with AD and MCI as well as cognitively normal (CN) individuals^35^. In this ADNI cohort, AD is defined based on the results of cognitive and clinical tests^35^. In addition to demographic and genetic information, ADNI includes longitudinal sampling of clinical metrics (Clinical Dementia Rating Sum of Boxes [CDRSB], Alzheimer’s Disease Assessment Scale-Cognitive Subscale [ADAS11 and ADAS13], Functional Activities Questionnaire [FAQ], Mini-Mental State Examination [MMSE]), blood biomarkers (plasma neurofilament light chain [NfL], plasma phosphorylated tau 217 [p-tau217], plasma amyloid-β40 and 42 and their ratio [Aβ40, Aβ42, Aβ42/Aβ40]), and blood metabolomics **(Figure 1A**). In this study, we used sparse longitudinal blood metabolomics from five different data modalities: 1) serum metabolites measured by flow injection analysis (FIA), 2) plasma lipidomics measured by liquid chromatography-mass spectrometry (LCMS), 3) serum metabolites measured by the Metabolon platform (METABOLON), 4) serum lipids and lipoproteins measured by nuclear magnetic resonance spectroscopy (NMR), and 5) serum metabolites measured by ultra-performance liquid chromatography mass spectrometry (UPLC). Each modality contained data which were largely generated from the same blood draws, providing matched multi-modal molecular measurements across time for 1,493 (639 AD; 339 MCI; 515 CN) unique individuals and 4,881 (2,172 AD; 1,058 MCI; 1651 CN) unique samples (**Supplementary Table 1**). Critically, samples were available before diagnosis of clinical AD for a number of participants (37 participants enrolled as CN and 344 participants enrolled as MCI that converted to AD during the course of the study).

### AD timelines inferred from sparse longitudinal blood metabolomics data model the unknown underlying timeline of AD

To conduct a genetic study of AD progression rate, we first inferred an AD timeline using our previously developed Timeline Inference Method (TIM)^34^. TIM uses a cross-validated machine learning approach to order longitudinal molecular samples in a large cohort in a way that maximizes order within each individual (**Figure 1B**). Briefly, we fit an elastic net regression model that uses molecular data to predict the relative timing of sample collection within each individual with AD (i.e., time since study entry). Because age at sampling is colinear with time since study entry, it is not possible to infer a timeline of AD while also adjusting for age at sampling. Instead, we ensure that the inferred timeline reflects AD progression rather than aging by using generalized contrastive principal component analysis (gcPCA)^36,37^ to construct features for timeline inference that minimize variation present in the CN group (e.g. differences due to age). Because our model does not force the order of samples within each individual to be correct, a unique advantage of our approach is that we can define an unbiased performance metric *τ*_timeline_. This performance metric measures how well our inferred timeline represents the true timeline underlying disease progression across all individuals with AD, ranging from 0 (random performance) to 1 (perfect performance)^34^. Each sample from subjects with AD, MCI or CN are then assigned a temporal location along the inferred AD timeline.

Using sparse longitudinal plasma metabolomics data from individuals with AD in ADNI described above, we inferred modality-specific timelines of AD progression. Across all modalities, our performance metric^34^ indicated that inferred timelines recapitulate temporal progression better than random (all *τ*_timelines_ > 0.34; **Figure 2A**), with serum metabolites (FIA) yielding the highest accuracy with *τ*_timeline_ = 0.58. To test whether these timelines captured true within-individual temporal progression rather than spurious structure in the data, null timelines were generated by randomly permuting timepoint labels within each individual. This procedure eliminated the signal in all modalities, yielding performance estimates near zero (modality-specific mean τ_timelineNulls_ ≤ [0.0000-0.0011], standard deviation = [0.023, 0.028], **Supplementary Figure 1B**) indicating that the inferred timelines depend on the true ordering of longitudinal samples and reflect meaningful temporal progression.

**Figure 2.**
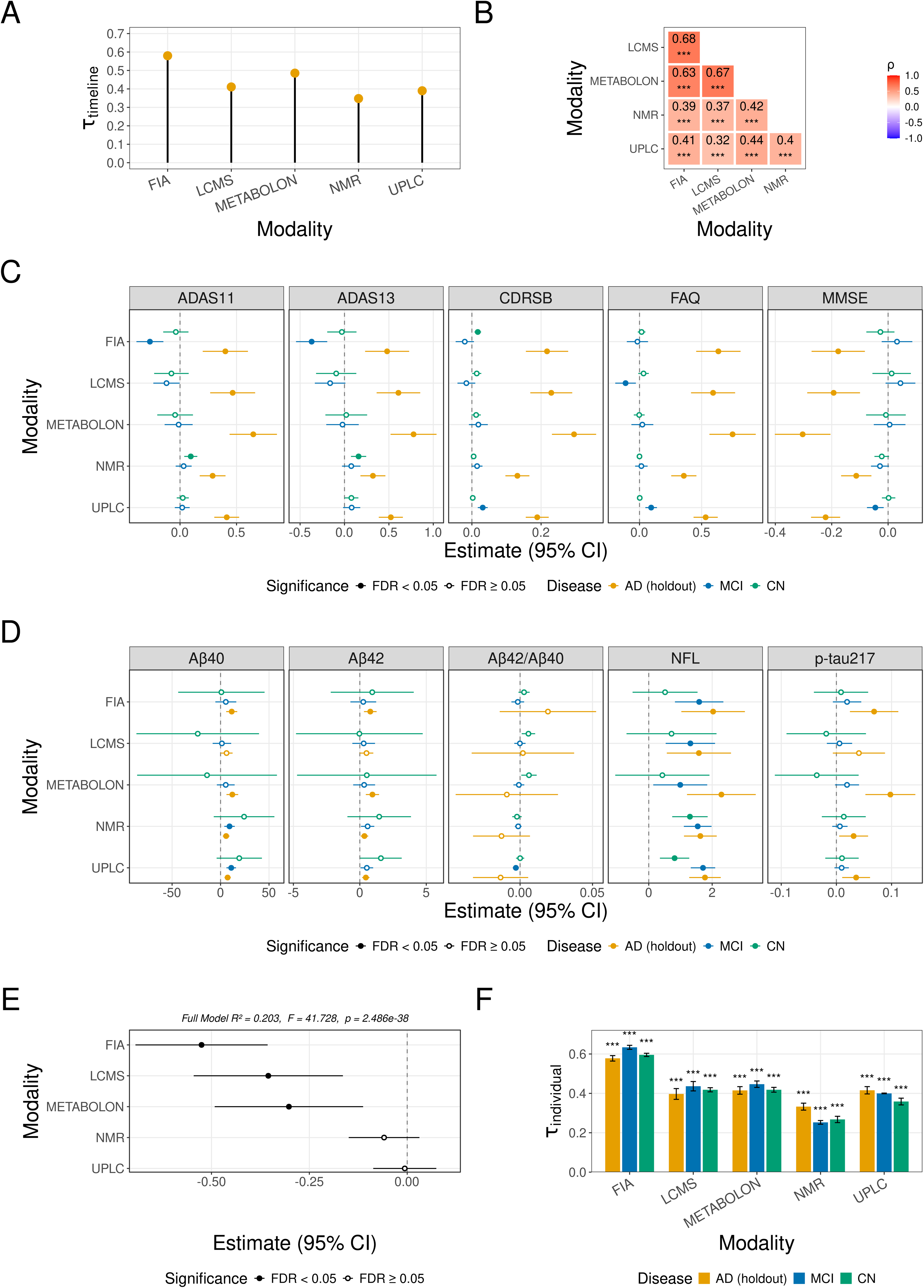
Inferred Timelines of AD Recapitulate Clinical Progression, Biomarker Accumulation, and the True Passage of Calendar Time. **(A)** *Lollipop Plot of Estimated Performance of AD Timeline Inference.* The x-axis is the modality of longitudinal metabolomics data used for timeline inference (FIA: serum metabolites measured by flow injection analysis, LCMS: plasma lipids measured by liquid chromatography-mass spectrometry, METABOLON: serum metabolites measured by metabolon platform, NMR: serum lipids and lipoproteins measured by nuclear magnetic resonance spectroscopy, and UPLC: serum metabolites measured by ultra-performance liquid chromatography mass spectrometry) and the y-axis the estimated performance (*τ_timeline_*) of AD timeline inference, ranging from 0 (random) to 1 (perfect). **(B)** *Heatmap of Correlations between Modality Specific Timelines in Individuals with AD.* Axes are the data modality used to infer the AD timeline, color scale (defined in the legend) and numbers in each box are the Spearman’s correlation (*ρ*) from each pairwise comparison, and asterisks denote significance as follows *** = FDR ≤ 8.39e-53. **(C & D)** *Forest Plots of the Association Between Cross-Validated (Methods) Temporal Location Along the AD Timelines and Clinical Outcomes* **(C)** *and Biomarkers of AD* **(D)**. The x-axis is the estimated effect size (and 95% confidence interval [CI]) of the associations between the temporal location along the AD timeline and the clinical metric or biomarker feature measured in blood plasma in pg/mL (facet title), and the y-axis the modality used for timeline inference. All associations control for age at sampling. Color denotes diagnostic group of held-out samples projected onto the AD timeline, fill denotes the FDR significance for the association, and both are defined in the legend. **(E)** *Forest Plot of the Association Between Cross-Validated (Methods) Temporal Location Along the AD Timelines and Age of Symptom Onset.* The x-axis is the estimated effect size (and 95% CI), and the y-axis the modality from which temporal locations for that predictor term is derived. Overall performance of the model with all terms is also shown above the plot. **(F)** *Bar Plot of Cross-Validated (Methods) Concordance with True Longitudinal Sample Ordering Within Individual (τ_individual_) Across Diagnostic Groups.* The x-axis is the data modality used to infer the AD timeline and the y-axis the concordance with true longitudinal sample ordering within individual (mean *τ_individual_* across 10 random seeds ± standard deviation). Color is the diagnostic group as defined in the legend, and asterisks the maximum FDR across seeds (*** = FDR ≤ 3.31 x 10^-11^).

Plasma metabolomics measurements in ADNI are largely derived from the same blood draws, providing an opportunity to test cross-modality consistency. We therefore assessed whether independently inferred timelines from distinct modalities converge on a shared underlying disease trajectory. Timelines inferred from each modality were highly concordant (Spearman’s ρ ≥ 0.32, FDR ≤ 8.39 x 10^-53^, **Figure 2B**). This concordance between distinct molecular feature spaces in largely overlapping samples provides strong evidence that the inferred AD timelines capture a shared and biologically meaningful disease trajectory, rather than modality-specific features or technical artifacts.

### Inferred AD timelines capture clinical and biological disease progression

Establishing that the inferred timelines track clinically meaningful disease progression is critical to demonstrating their validity for clinical and biological analyses, including genetic studies. Accordingly, we assessed whether temporal location recapitulates clinical progression in three-fold cross-validation (Methods). Across all modality-specific timelines, temporal location was significantly associated with all tested measures of clinical severity in AD individuals while controlling for age at sampling (**Figure 2C**). Specifically, later temporal locations were significantly associated with worse clinical severity (increased CDRSB, ADAS11, ADAS13, and FAQ scores; and decreased MMSE scores, **Figure 2C**) consistent with expected disease progression. In contrast, samples from MCI and CN individuals projected onto the same AD timelines were generally not significantly associated with clinical severity and not consistently aligned with the expected direction of disease progression (**Figure 2C**). Together, these results demonstrate that the inferred timelines recapitulate the clinical progression of AD in an out-of-sample setting and in a disease-specific manner, distinguishing AD-related progression from variation observed in MCI and CN individuals.

We next investigated whether temporal location along the inferred AD timelines was associated with established plasma biomarkers of AD, again controlling for age at sampling. These analytes are mostly used as diagnostic markers of AD, but have also been suggested to track pathophysiological progression^38–42^. Significant associations with temporal location along the inferred AD timeline were observed for plasma Aβ42 and p-tau217 (**Figure 2D, Supplementary Table 2**). Interestingly, nearly no associations were observed for Aβ42/Aβ40 ratio despite its established value for identifying AD pathology. These findings indicate that our timelines capture a component of AD progression that is overlapping yet distinct from classical biomarkers that define whether AD pathology is present. In contrast, later temporal location was more consistently associated with higher plasma Aβ40 and, most prominently, higher plasma NfL (**Figure 2D**). Significant associations with Aβ40 were observed in four of five modalities in AD, two in MCI, and none in CN individuals (**Figure 2D**). Plasma NfL exhibited the clearest and most consistent relationship with temporal location, with higher NfL generally associated with later temporal location across AD, MCI, and CN groups (**Figure 2D**). Together, these results further support the biological validity of the inferred timelines while also highlighting an important conceptual distinction: diagnostic biomarkers of AD may only capture a subset of the true progression of the disease, and plasma NfL may better track temporal progression than core AD diagnostic biomarkers.

### Temporal location on inferred AD timeline predicts age of symptom onset

Although the inferred AD timelines recapitulate multiple aspects of AD biology, an important question is whether they capture the true passage of calendar time. In AD, the true onset of disease is not directly observable, and the onset of disease symptoms occurs years to decades after pathological processes begin and is itself subject to substantial variability^43,44^. Thus, symptom onset date provides only a noisy and imperfect proxy for disease timing. Nevertheless, as the only available temporal reference in ADNI, we assessed whether temporal location along the inferred timelines was associated with time relative to AD symptom onset date. Using a linear model with temporal locations from all five modality-specific timelines as predictors, we found a significant association with time since symptom onset (r^2^ = 0.20, p = 2.49 × 10^-38^; **Figure 2E**). Despite the noise present in symptom-based temporal reference, this result indicates that the inferred timelines capture a meaningful temporal signal aligned with disease progression in calendar time.

### Longitudinal sample ordering from MCI and CN individuals is recapitulated along AD timelines

Finally, we assessed whether the inferred AD timelines preserve within-individual temporal ordering in both non-AD and AD individuals. Using the same cross-validated framework, we performed an exact statistical test (block *τ* test) to assess whether pairs of samples from the same individual were correctly ordered along the inferred timeline in AD, MCI and CN groups^34^. Sample pairs were correctly ordered significantly above chance in all groups (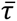_individual_ range of [0.25-0.63], **Figure 2F**), indicating that the inferred AD timelines capture consistent temporal structure within individuals beyond the AD population. These results contrast with the disease-specific associations observed with clinical measures, which were largely restricted to AD. To assess whether the inferred AD timelines recapitulate disease progression rather than aging, we tested whether the temporal location along these timelines was correlated with age at sampling across individuals. The magnitude of correlation (Kendall’s *τ*) for the temporal location along inferred AD timelines with age at sampling (Kendall’s *τ* range of [0.07-0.22], **Supplementary Table 2**) was substantially lower than *τ*_timeline_. Because *τ*_timeline_ is directly comparable to Kendall’s *τ*^34^, this makes it unlikely that the inferred timeline of AD is simply a representation of processes of aging rather than those of disease progression. Together, these findings suggest that while the inferred temporal locations are specific to AD in their clinical relevance, the AD timelines capture a broader temporal axis that is present in individuals regardless of diagnosis. This justifies inclusion of MCI and CN individuals in genetic studies based on these timelines, and additionally supports a paradigm of AD risk that treats progression along the AD timeline and potential to develop clinical AD as at least partially distinct processes.

### Individual AD timeline progression rates are concordant across data modalities

Having established that the inferred AD timelines represent disease progression, we next sought to define an individual-level quantitative trait suitable for genetic analysis that characterizes variability in individual rates of progression along these timelines. Our approach infers a timeline of disease without assuming a fixed rate of progression, allowing individuals to advance along the AD timeline at different rates relative to calendar time, thereby enabling estimation of individual-specific timeline progression rates. Specifically, we define individual timeline progression rates as the ratio between the change in estimated temporal location and the true passage of time in that period, estimated with linear regression (**Figure 3A**). We then assessed whether these estimated timeline progression rates capture meaningful variation in disease dynamics.

**Figure 3.**
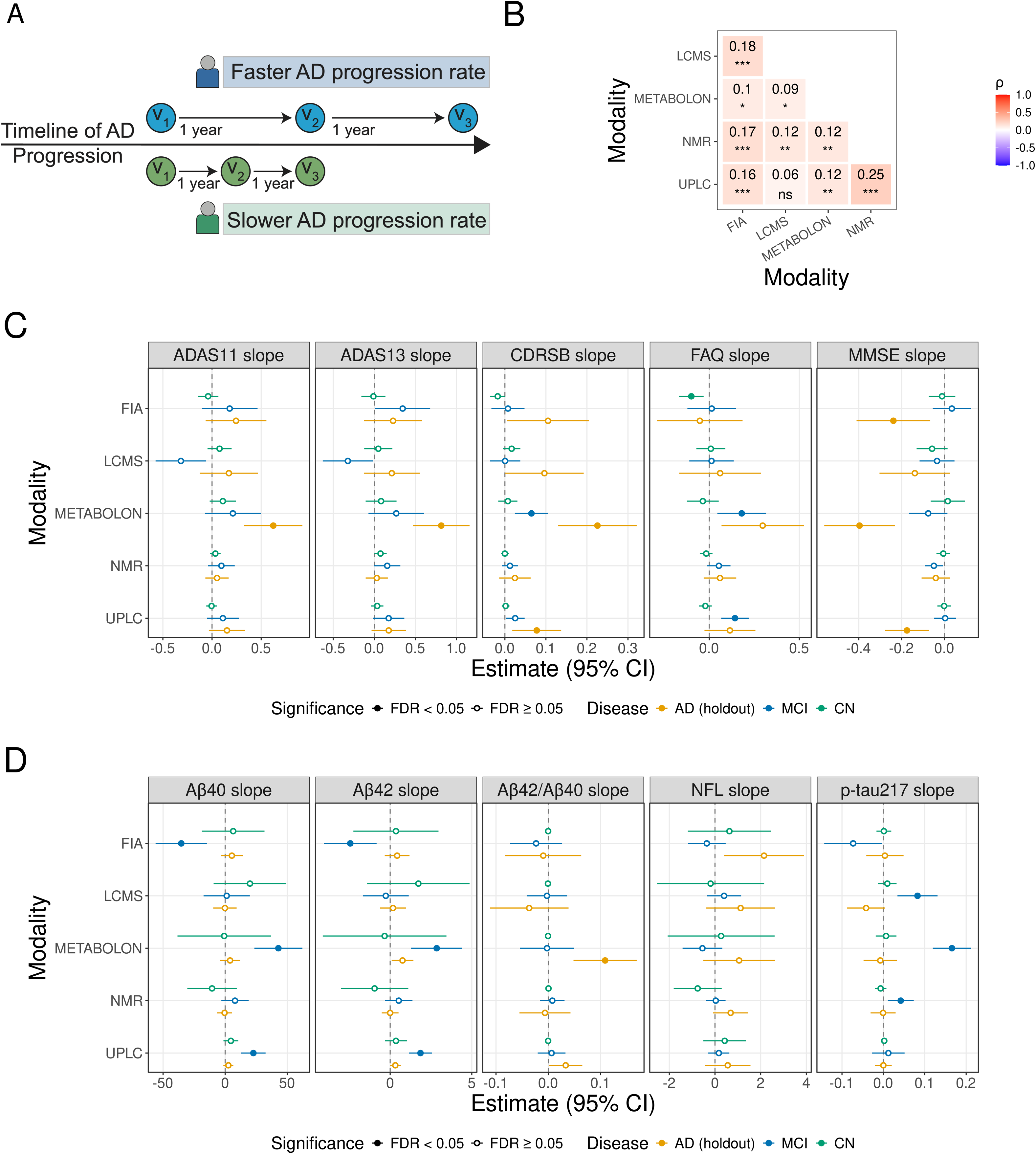
AD Timeline Progression Rates Capture Clinical and Biological Disease Progression. **(A)** Schematics of AD Timeline Progression Rate Across Faster (blue) and Slower (green) Progressors. **(B)** Heatmap of Correlations between Modality Specific AD Timeline Progression Rates in Individuals with AD. Axes are the data modality used to infer the AD timeline, color scale (defined in the legend) and numbers in each box are the Spearman’s correlation (*ρ*) from each pairwise comparison, and asterisks denote significance as follows *** = FDR ≤ 0.001, ** = FDR ≤ 0.01, * = FDR ≤ 0.05, ns = not significant. **(C & D)** Forest Plots of the Association Between Cross-Validated (Methods) AD Timeline Progression Rate and Clinical Outcomes **(C)** and Biomarkers of AD **(D)**. The x-axis is the estimated effect size (and 95% CI) of the associations between the AD timeline progression rate and the rate of change of clinical metric or biomarker feature (facet title) over time (calendar years), and the y-axis the modality used for timeline inference. All associations control for age at baseline. Color denotes diagnostic group of held-out samples projected onto the AD timeline, fill denotes the FDR significance for the association, and both are defined in the legend.

We first assessed the consistency of individual-specific AD timeline progression rates across modality-specific timelines. Despite being derived from distinct molecular feature spaces, AD timeline progression rates were generally significantly positively correlated in AD (for 9/10 pairwise comparisons FDR ≤ 0.05 with Spearman’s ρ range of [0.09-0.25], **Figure 3B**) and across other disease groups (**Supplementary Figure 2A, 2B),** indicating that they capture a shared underlying signal. However, the strength of these correlations (Spearman’s ρ ≤ 0.25, **Figure 3B)** was consistently weaker than the correlations observed for temporal location along the same timelines (Spearman’s ρ ≥ 0.32, **Figure 2B**). This lower correlation is expected, as timeline progression rates are a second-order estimate derived from estimated temporal locations and are therefore more sensitive both to error in timeline inference and noise in longitudinal sampling. Together, these results indicate that AD timeline progression rates exhibit cross-modality consistency while also having their own distinct properties.

### AD timeline progression rates capture clinical and biological disease progression

We next assessed whether AD timeline progression rates capture aspects of disease dynamics represented by rates of clinical progression. A progression rate was defined for each clinical measure analogously to timeline progression rate, as the rate of change of the clinical severity over time within individuals. Using the same cross-validated framework as presented above, we tested whether AD timeline progression rates were associated with the clinical progression rates for battery scores across diagnostic groups, adjusted for age at initial sampling (**Figure 3C**).

Estimated AD timeline progression rates were significantly associated with rates of progression of several clinical measures in the AD group, with effects consistently observed in the expected direction (i.e., faster progression along the AD timeline corresponding to faster progression in clinical scores). Notably, the magnitude of association varied across data modalities and clinical phenotypes, indicating that molecular profiles may capture non-overlapping signals. In MCI and CN individuals, associations were generally weaker, although some trends in the expected direction were also observed. Consistent with these results, joint tests of association between all modality-specific AD timeline progression rates and clinical decline (F-test) demonstrated significant relationships with all clinical measures except FAQ in the AD group, with weaker or non-significant effects in MCI and CN (**Supplementary Figure 2C**). Together, these findings indicate that AD timeline progression rates capture meaningful variation in the clinically-derived rate of decline, particularly in individuals with AD, supporting their clinical relevance.

We next assessed whether AD timeline progression rates were associated with rates of change in plasma levels of known AD biomarkers. Biomarker progression rates were defined in the same way as clinical progression rates, as the slope of biomarker levels over time within individuals (**Figure 3D**). In contrast to clinical measures, we observed limited and inconsistent associations between AD timeline progression rates and biomarker progression rates across modalities and diagnostic groups. While some associations were detected for plasma Aβ40, Aβ42, and the Aβ42/Aβ40 ratio in specific modality-group combinations, these effects were inconsistent in direction and variable in magnitude. More broadly, no single biomarker or modality provided a robust or reproducible association with AD timeline progression rate. Consistent with these observations, joint tests of association (F-test) generally showed inconsistent relationships (**Supplementary Figure 2D**). This is consistent with the established model of progression of classical AD biomarkers, in which they change non-linearly along a sigmoid curve over the temporal course of the disease and are thus not expected to progress linearly over time^45–48^. Together, these results indicate that, while commonly measured plasma biomarkers of AD track pathophysiological progression, the rate of that progression cannot be straightforwardly estimated from them, underscoring the limitations of current biomarkers in capturing complete disease dynamics.

### GWAS of AD timeline progression rate reveals genetic signals distinct from disease risk

Having established that AD timeline progression rates capture biologically meaningful variation in disease dynamics across individuals, we can use these quantitative traits to characterize the genetic architecture of AD progression. Given the results presented above, we hypothesize that the total genetic risk for AD may comprise at least two distinct components: the risk for the potential of developing the disease independently of the passage of time (i.e. risk for AD potential) and the risk for progression along the disease timeline (i.e. risk for rate of AD progression), both of which contribute to ascertainment of case-control status. AD timeline progression rate is an individual-specific quantitative trait that captures risk for AD progression rate directly. Therefore, we performed common variant (minor allele frequency [MAF] > 0.01) GWAS of AD timeline progression rate for each data modality to identify variants and genes associated specifically with rate of AD progression.

Because temporal ordering was robust across diagnostic groups, (**Figure 2F**), we performed GWAS of AD timeline progression rate separately in AD, MCI, and CN cohorts, followed by meta-analysis across groups to maximize power. GWAS were well-controlled, as evidenced by quantile-quantile plots showing minimal inflation (all λ ≤0.99, **Supplementary Figure 3A-3E**), and yielded multiple genome-wide significant and suggestive associations (**Figure, 4A-4E**; **Table 1**). Across data modalities, we identified three genome-wide significant loci (P ≤ 5 x 10^-8^), two from plasma lipidomics (LCMS) and one from serum lipids and lipoproteins (NMR) and many suggestive associations across modalities (**Figure, 4A-4E**; **Table 1**). As expected at our sample size, we were unable to obtain a direct estimate of heritability using methods such as LD score regression, LDAK, or GREML (1,341 individuals with both genetic data and AD timeline progression rate estimates compared to a minimum recommended sample size of 5,000-10,000 for all these methods)^49–54^. Nevertheless, despite the lack of a direct measurement of heritability, these GWAS results indicate that genetic signals associated with AD timeline progression rate can be detected, supporting that this phenotype is under genetic control.

**Table 1.**
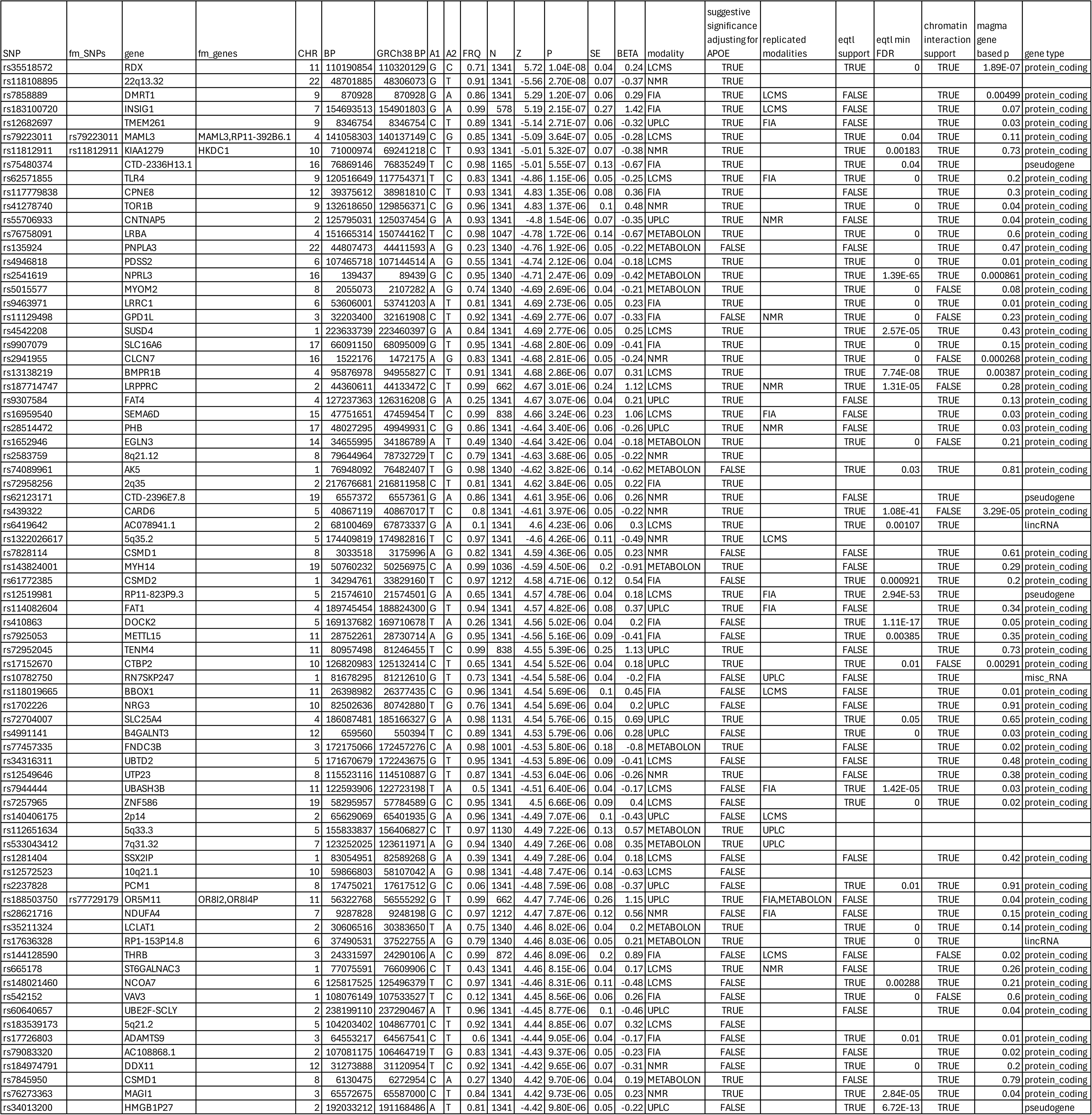
Summary of Association Results with AD Timeline Progression Rate. Each row is defined by the combination of locus and data modality used for timeline inference and is included in this table when an association with AD timeline progression rate is observed with P ≤ 1 x 10^-5^. The columns of the table contain the following information for GRCh37 unless otherwise specified: lead rsID (SNP); finemapped rsIDs (fm_SNPs); prioritized genes from FUMA (gene); genes mapped to fine-mapped SNPs (fm_genes); genomic location (CHR, BP); base pair location in GRCh38 (GRCh38_BP); reference allele (A1); alternate (effect) allele (A2); effect allele frequency in assessed cohort (FRQ); effective sample size (N); z score for association (Z); p value for association (P); standard error for association (SE); effect size for association (BETA); data modality where association is observed (modality); whether association is replicated at P ≤ 1 x 10^-5^ with concordant direction of effect while controlling for *APOE* genotype (suggestive_significance_with_APOE_genotype_covars); and modalities where association is replicated at P ≤ 0.05 with concordant direction of effect (replicated_modalities). The table also contains columns for prioritized mapped gene support: whether eQTL association was observed (eqtl_support); minimum FDR for eQTL association from FUMA (eqtl_min_FDR); chromatin interaction support from FUMA (chromatin_interaction_support); gene-based association from MAGMA (magma_gene_based_p); and gene biotype (gene_type).

Because our inferred AD timeline progression rate is not directly analogous to any phenotype previously tested for genetic associations in other cohorts, direct replication in independent datasets is not possible. We therefore sought indirect validation by testing whether loci associated with AD timeline progression rate were replicated in existing GWAS of other AD-related traits. These included GWAS of observed AD case-control status, GWAS of AD family history, and GWAS of AD biomarkers and pathology. Notably, observed case-control status is not independent of progression dynamics, as individuals with faster progression are more likely to be ascertained as AD cases. Meanwhile, biomarker GWAS are relevant given the observed relationships between biomarkers and AD timeline progression rate. Across modalities, loci associated with AD timeline progression rate (P ≤ 5 x 10^-5^) were replicated (concordant direction of effect and P ≤ 0.05) in multiple external GWAS datasets for AD and AD related phenotypes (**Figure 4F, Supplementary Table 4**), supporting the validity of the identified signals. In total, across all 5 modalities, more loci than expected by random chance were replicated in 20 tests in 16 external GWAS (one-sided binomial test, FWER adjusted P ≤ 0.05), with the best replication for serum metabolite (Metabolon) and dementia^55^ (29% replication rate, FWER P = 1.57 x 10^-78^, **Figure 4F, Supplementary Table 4**). Replication patterns varied across data modalities, with AD timeline progression rates derived from specific molecular profiles showing stronger enrichment in certain AD and AD-related phenotypes than others (**Figure 4F**; **Supplementary Table 4**), supporting that different modalities may capture partially distinct aspects of disease progression.

**Figure 4.**
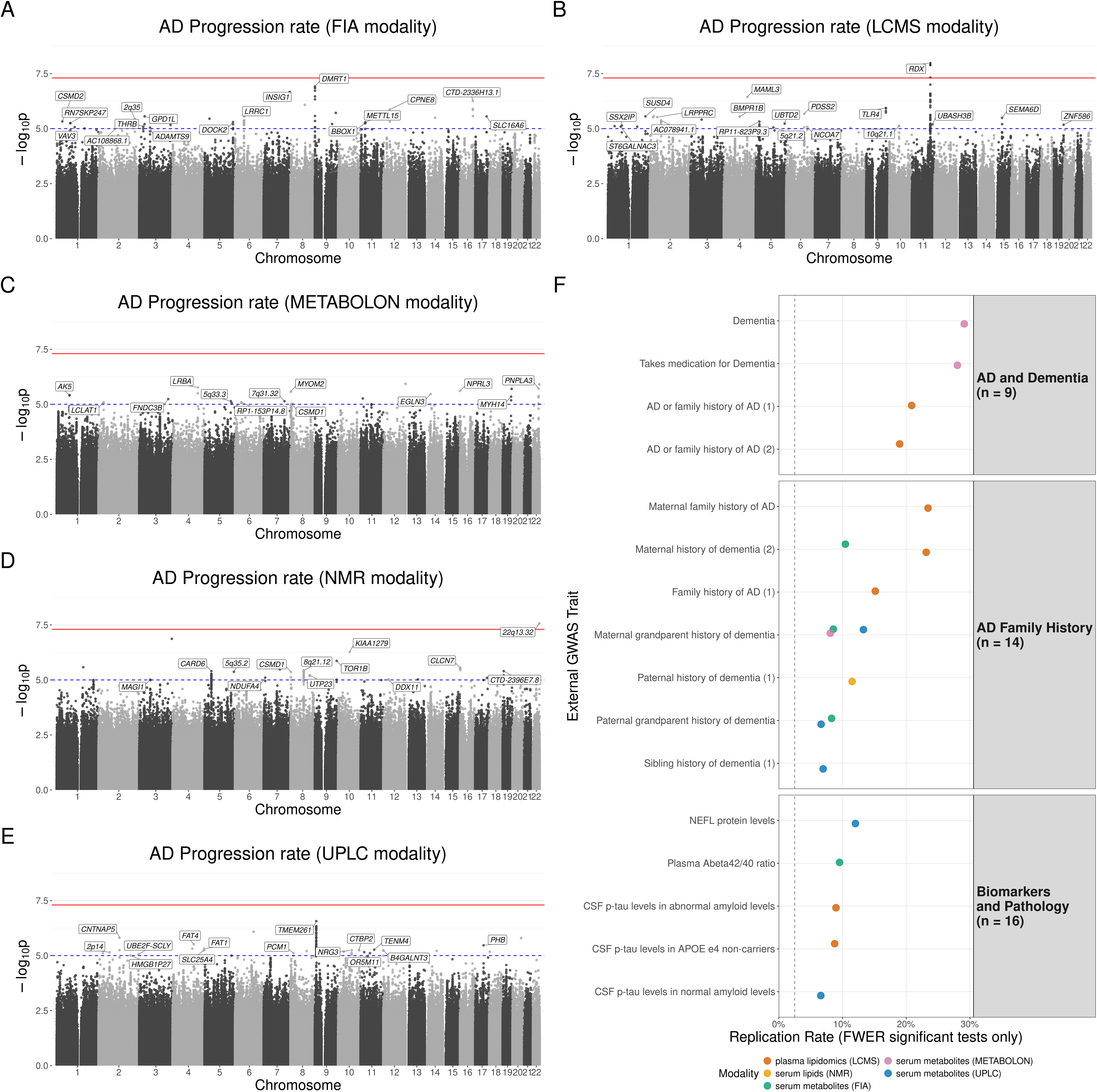
AD Timeline Progression Rate is Associated with Novel Genetic Loci. **(A-E)** *Manhattan Plots of GWAS Results for AD Timeline Progression Rates.* The x- and y-axes are genomic locations and -log_10_(p-values) for variants (points) tested for association with AD timeline progression rate. The red and blue lines show genome-wide significance threshold (P ≤ 5 x 10^-8^) and suggestive significance threshold (P ≤ 1 x 10^-5^). Mapped genes summarized in Table 1 are indicated on the plots. Data modality used for timeline inference: FIA **(A)**, LCMS **(B)**, METABOLON **(C)**, NMR **(D)**, and UPLC **(E)**. **(F)** *Replication of Suggestive Signals (P ≤ 5 x 10^-5^) in External GWASs of AD Related Phenotypes.* The x-axis is the replication rate among Bonferroni-corrected significant tests from a one-sided binomial test (concordant direction of effect and P ≤ 0.05 in external GWAS) and the y-axis is the external study phenotype assessed. The vertical dashed line shows expected null replication rate (2.5%). Facets represent groupings into major phenotypic themes (n = number of external studies tested from that group). Color is the data modality used for timeline inference and is defined in the legend.

To further characterize loci associated with AD timeline progression rate, we performed statistical and functional fine-mapping using SAFFARI^56^, identifying putative causal variants for 11 loci (**Table 1**). We also mapped associated loci to genes using FUMA through multiple complementary approaches, including gene-based association testing (MAGMA)^57,58^, expression quantitative trait loci (eQTL) mapping, and chromatin interaction mapping. This integrative strategy prioritized candidate genes for 67 timeline progression-associated loci (**Table 1**; full list of prioritized and non-prioritized candidate genes in **Supplementary Table 5**). Among these, *RDX* (identified in the plasma lipidomics (LCMS) analysis) emerged as the most strongly supported candidate gene, with convergent evidence from all three gene mapping approaches. Other genes included *DOCK2*, *KIAA1279*, *TLR4*, and *SEMA6D* (**Table 1**). Notably, many of these genes have not previously been implicated in AD genetics. This highlights the potential of progression-based genetic analyses to reveal aspects of disease biology not captured by traditional observed case-control studies, in addition to identifying several novel loci associated with AD timeline progression rate.

### AD timeline progression rate genetics are independent of *APOE* genotype

Given the central role of *APOE* in AD genetics^18–21^, we next assessed whether the identified genetic associations with AD timeline progression rate were dependent on *APOE* genotype. Indeed, many AD-associated genetic signals are partially or fully driven by *APOE* genotype^59,60^. Notably, we did not observe a prominent signal at the *APOE* locus in any of our GWAS (**Figure 4A-4E**; **Table 1**), prompting us to explicitly test for potential dependence. To this end, we repeated all GWAS analyses adjusting for *APOE* genotype. Including the number of ε2 and ε4 alleles carried by each individual as additional covariates had minimal impact on the results, with near-identical association statistics with and without *APOE* genotype adjustment observed across all modalities (**Table 1**; **Supplementary Figure 3F-3J; Supplementary Table 3-4**). This was reflected in strong concordance of Z-scores between models with and without *APOE* adjustment (all ρ > 0.97; **Supplementary Figure 3F-3J**), as well as consistent p-value distributions across phenotypes (all ρ > 0.91; **Supplementary Figure 3F-3J**). To exclude the possibility that these results were driven by linkage disequilibrium (LD) structure, we performed LD pruning and compared effect sizes between models with and without *APOE* adjustment. In all cases, effect sizes remained highly correlated (Spearman ρ > 0.97, **Supplementary Figure, 3K, Supplementary Table 4**), confirming independence from *APOE*. Together, these results demonstrate that the genetic architecture of AD timeline progression rate is largely independent of *APOE*, distinguishing it from traditional GWAS of observed AD case-control status and suggesting that progression-based analyses capture complementary aspects of disease biology.

### Mendelian Randomization (MR) analysis confirms AD timeline progression rate is causal to clinical and molecular progression

The results above demonstrate that both temporal location and progression rate along the AD timeline have strong, significant relationships with multiple clinical measures of AD and disease progression, as well as weaker relationships with many plasma biomarkers. Successful discovery of genetic markers associated with AD timeline progression rate allows us to use MR analysis to validate the causal structure of these relationships. MR analyses were first performed on measures of progression rate of clinical severity to assess whether AD timeline progression rate is causal to these measures. Briefly, we constructed genetic risk scores for modality-specific AD timeline progression rates, which were then used as instruments to perform one-sample MR using the two-stage least squares (2SLS) method^61^, treating estimated AD timeline progression rate as the exposure and five measures of clinical severity (CDRSB, ADAS11, ADAS13, MMSE, and FAQ) as the outcome. Across all five measures of severity and four of five modalities, higher genetic risk of AD progression was significantly associated with faster progression of clinical severity over time (**Figure 5A**). This confirms that our individual AD timeline progression rate is estimating a clinically relevant property that is causal to progression of clinical severity. In addition to testing rate of change of clinical severity over time, clinical severity at study enrollment, which is expected to be more severe in faster-progressing individuals due to progression occurring before enrollment, was also assessed. We found significant causal relationships between all five modality-specific AD timeline progression rates and all five measures of clinical severity at enrollment, further confirming the clinical relevance of estimated AD timeline progression rate (**Figure 5B**).

**Figure 5.**
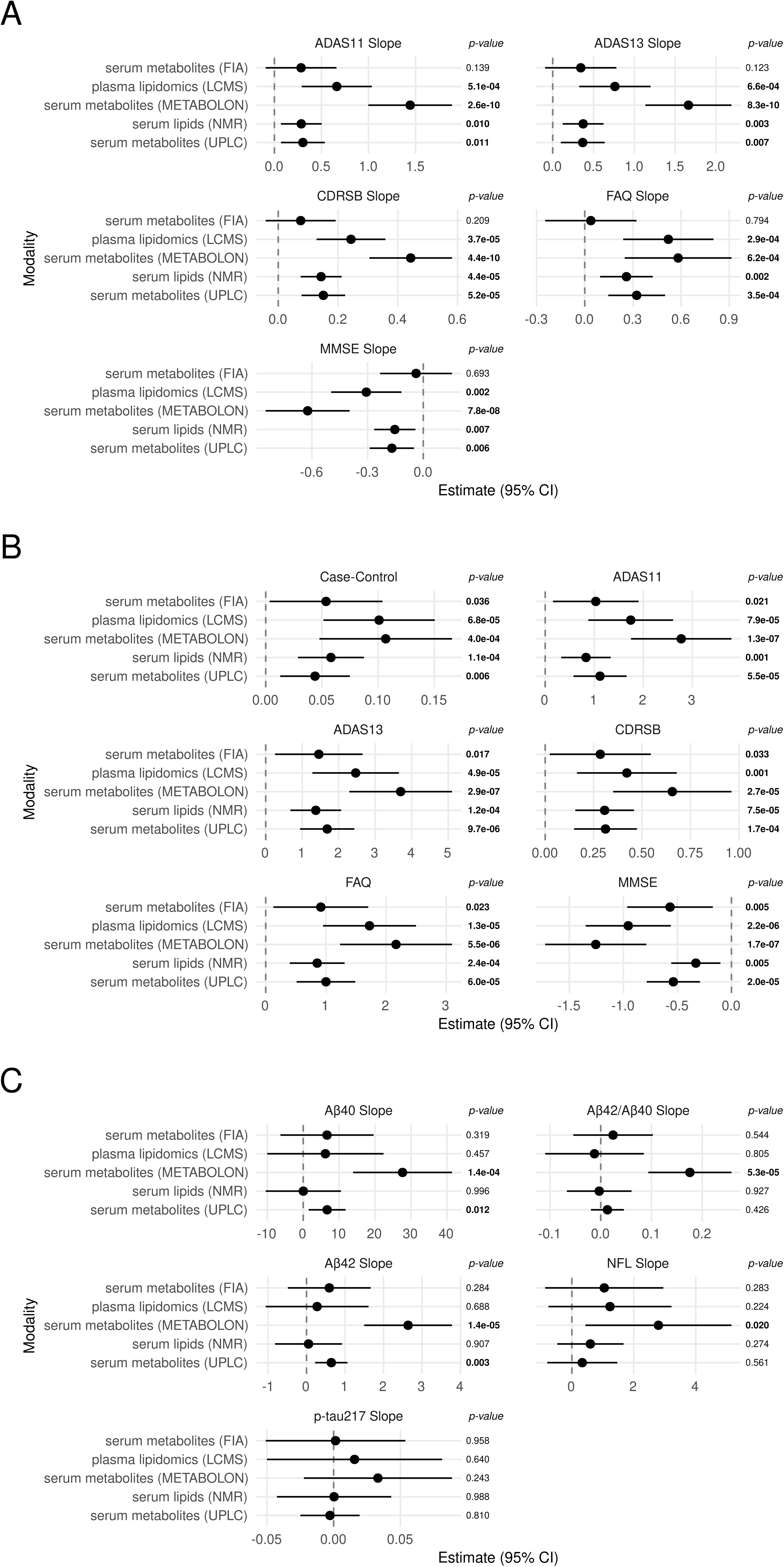
AD Timeline Progression Rate is Causally Linked to Clinical and Biological Disease Progression. *Forest Plots of the Results of One Sample MR Using AD Timeline Progression Rate as the Exposure and AD Traits as Outcomes for: Progression of Clinical Outcomes **(A)**, Clinical Outcomes at Baseline **(B)**, and Progression of Biomarkers of AD **(C)**.* The x-axis is the estimated effect size (and 95% CI) of the causal relationship between the AD timeline progression rate and the assessed outcome (facet title), and the y-axis the data modality used for timeline inference. P-values for each association are shown on the right of each forest plot. Case-control associations are in log(OR) units. Units for all other associations are either those of untransformed scores at baseline, or rate of change of clinical metric or biomarker feature (facet title) over time (calendar years).

Next, the same MR analyses were performed on measures of progression rate of plasma biomarkers. As observed for the AD timeline progression rate-plasma biomarker analyses described above (**Figures 2D and 3D**), results were more varied between modalities and less significant. Similar to the observations from the biomarker association analyses (**Figure 3D**), AD timeline progression rate was not identified as causally linked to the rates of change over time of the diagnostic biomarker p-tau217 (**Figure 5C**). Although this does not rule out a potential causal relationship that could be captured by a different model, these results reinforce that AD timeline progression rate is not necessarily straightforwardly related to the rate of change of diagnostic biomarkers over time (**Figure 3C**). However, unlike in this previous analysis, MR showed a directionally consistent relationship between AD timeline progression rate and rates of change of Aβ40, Aβ42, and NfL over time, including significant (P ≤ 0.05) associations with up to two of the three serum metabolite modalities (METABOLON and UPLC) (**Figure 5C**). These results further implicate individual AD timeline progression rates as a meaningful biological measure that is causally related to pathophysiological progression as measured by both clinical presentation and plasma biomarkers.

Finally, MR was performed to assess the relationship between AD timeline progression rate and observed case-control status. Across all five modalities, a significant causal effect of AD timeline progression rate on case-control status was observed (**Figure 5B**). This relationship is consistent with case ascertainment dynamics, as individuals must reach a sufficient level of disease progression to be classified as AD cases. Importantly, this signal is not driven by explicit case-control comparisons in our framework: timelines were inferred using only AD cases, and GWAS of AD timeline progression rate were performed within diagnostic groups and meta-analyzed across AD, MCI, and CN cohorts. As such, neither temporal location nor genetic associations were informed by case-control status. The strong causal relationship observed here therefore provides independent support that AD timeline progression rate is a key determinant of observed AD case-control status.

### A multidimensional genetic paradigm of AD risk integrating risk for AD potential and risk for AD progression rate

Genetic risk of AD is typically modeled using a liability threshold framework, in which genetic loci contribute to an individual’s total “liability” or “susceptibility” alongside other factors, including lifestyle, environmental exposures, and aging^12,13,62^. Individuals develop AD when their total susceptibility exceeds a threshold and remain unaffected if their total susceptibility never crosses that threshold. Under this model, case-control GWAS is conceptualized as a method to detect genetic contributors to this underlying quantitative trait of total susceptibility (“genetic susceptibility”), which is possible because total susceptibility determines case-control status^63–65^. Our results show that loci controlling AD timeline progression rate replicate in AD case-control GWAS and are causally linked to both AD pathophysiology and case-control status. In the context of the liability threshold model, these results imply that progression rate contributes to AD genetic susceptibility. However, the contribution of AD timeline progression rate to AD genetic susceptibility is independent of the major risk locus *APOE*, with no detectable association in the *APOE* locus and no change in genetic associations after controlling for *APOE* genotype, suggesting that the biology underlying progression is at least partially distinct from that captured by case-control GWAS.

To resolve this discrepancy in a manner that allows AD progression rate to be under separate genetic control and total susceptibility for AD to increase with age, we propose a multidimensional, longitudinal extension of the standard liability threshold model. In this model, total AD risk arises from the combination of two distinct components: “risk for AD potential,” representing a risk component that controls the risk of developing the disease given infinite lifetime, and “risk for rate of AD progression,” representing a component that influences the temporal course of the disease, as measured by our GWAS for AD timeline progression rate (**Figure 6A**). Genetic susceptibility to AD as measured by case-control GWAS depends on both of these factors, although there are many possibilities for the precise functional form of this combination (Methods). Thus, we hypothesize that loci associated with AD case-control status and loci associated with AD timeline progression rate should be independently predictive of AD related phenotypes such as symptomatic age at onset (AAO).

**Figure 6.**
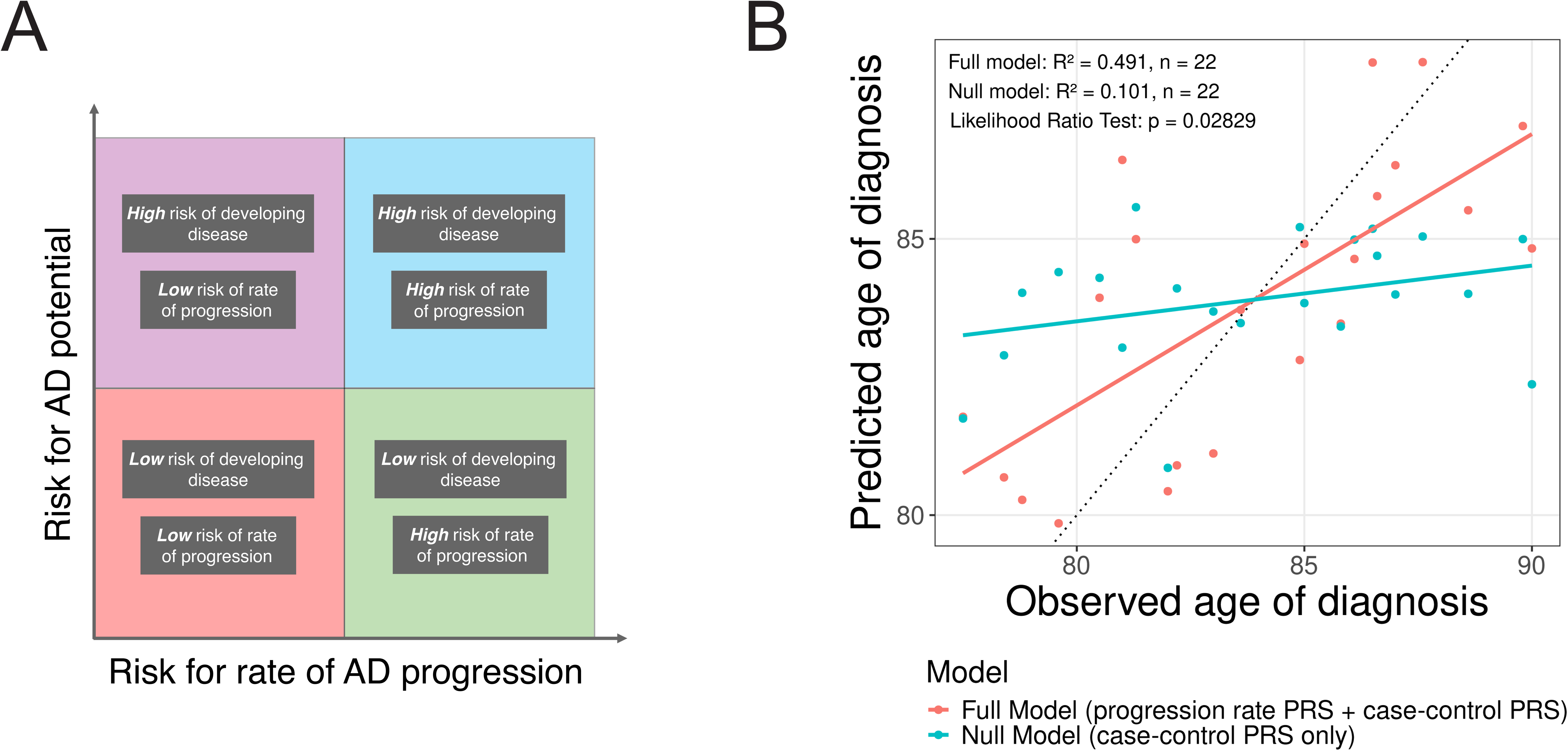
A Novel Paradigm for AD Genetic Risk. **(A)** Schematic for Total AD Genetic Risk as a Function of Risk for Rate of AD Progression (x-axis) and Risk for AD Potential (y-axis). Four example decompositions of total AD genetic risk are depicted in distinct quadrants. **(B)** Scatterplot of the Relationship Between Observed Age of Diagnosis (x-axis) and Predicted Age of Diagnosis (y-axis) Using AD Polygenic Risk Score (PRS) Only (Null Model), or AD PRS and AD Timeline Progression Rate PRS (Full Model). Color is the model used to predict age of diagnosis. Performance for each model (linear models) and statistic for the comparison of the full model relative to null model (Likelihood Ratio Test) are shown.

To test this hypothesis, we constructed polygenic risk scores (PRS) for both observed AD case-control status^13^ and AD timeline progression rate, and evaluated their ability to predict AAO in an independent subset of the ADNI data. To get the most accurate definition of AAO, this analysis is restricted to individuals who received an AD diagnosis during the course of the study. Consistent with our hypothesis, combining observed case-control PRS with timeline progression rate PRS significantly improved prediction of AAO compared to case-control score alone (case-control PRS only R^2^ = 0.101, case-control and AD timeline progression rate PRS R^2^ = 0.491; p = 0.0283, likelihood ratio test; **Figure 6B, Supplementary Table 4**). This result was replicated in the AD sequencing project (ADSP) data, where the combined observed case-control PRS and timeline progression rate PRS significantly improved prediction of reported AAO compared to observed case-control PRS alone (case-control PRS only R^2^ = 0.0337, case-control and AD timeline progression rate PRS R^2^ = 0.0359; p = 0.00371, likelihood ratio test; **Supplementary Table 4**). This result validates the notion that risk for AD potential and risk for rate of AD progression capture complementary genetic contributions to total AD susceptibility, providing a more complete representation of disease risk than either component alone and highlighting the importance of modeling disease dynamics to uncover genetic influences not captured by traditional approaches.

## Discussion

Significant effort has been exerted to integrate longitudinal modeling to better understand the genetic basis of diseases, including AD^29–32,66,67^. In this study, we applied our previously developed timeline inference approach to sparse longitudinal molecular data to characterize the genetic architecture of AD progression rate. The timeline inference framework enables estimation of both a temporal location for any sample in the same modality of data and a progression rate for any individual with multiple samples. We showed that both AD timeline temporal location and AD timeline progression rate align strongly with progression of clinical severity, a remarkable finding given the relative lack of accepted biomarkers for AD progression^38–42^. The same timeline-derived variables were also associated with diagnostic plasma biomarkers of AD, although associations were generally weaker. GWAS were performed for inferred AD timeline progression rates and several loci identified as potentially implicated in this new phenotype. Replication in existing published GWAS of AD-related phenotypes and MR analyses demonstrated that this new phenotype is a biologically meaningful measure of AD progression with distinct genetic architecture independent of established risk loci such as *APOE*, while still being closely related to existing GWAS of observed AD case-control status. Based on these findings and previous postulates^33^, we propose a new multidimensional paradigm of AD genetic risk in which total disease risk can be decomposed into two components: one influencing potential for disease, and one influencing rate of disease progression.

These inferred AD timelines recapitulate observed clinical and temporal features of disease progression. Other approaches have been proposed to reconstruct disease timelines from longitudinal biomarker data^68–70^. Many rely on time-to-event frameworks^71,72^ or proxies such as expected year of onset^68^, which depend on poorly defined reference events along a disease timeline that is not directly observable. In addition, these approaches are unable to scale to large multidimensional data, thereby often incorporating only a limited set of disease biomarkers, restricting their ability to capture the full complexity of AD progression^68^. Other approaches, such as pseudotime methods^73–75^, are designed for analyses of cross-sectional data, and as such cannot distinguish within-individual signals such as disease progression from inter-individual heterogeneity. As a result, these approaches cannot guarantee that any learned timeline is representative of true temporal variation rather than other axes of variation dominating the data. A common feature of all these approaches, previously described as the major remaining hurdle in that field, is that there is no way to estimate the accuracy of the inferred timelines^68^. Our data-driven timeline inference approach^34^ overcomes these limitations by changing how the temporal information contained in the longitudinal data is used. Our approach does not require predefined reference events, is not limited in the number of features it can use, does not assume equal progression rates between individuals, and most importantly estimates the performance of the timeline reconstruction directly from the data. Critically, these properties enable our approach to quantify disease progression rate as an individual-level trait relative to an individual’s temporal location along the reconstructed timeline, without which this study would not be possible.

While several biomarkers exist for diagnosing AD, such as plasma Aβ and p-tau measures, there are few established biomarkers of disease progression^38–42^. Our inferred temporal locations, derived from blood metabolites including lipids and peptides, were strong predictors of clinical severity over time, and MR analyses confirm that the timeline estimates a genuine biological property that is causally upstream of changes in clinical severity. In contrast, canonical AD biomarkers (e.g., Aβ42/Aβ40, p-tau217) show weaker associations with temporal location, but other analytes not typically used by themselves for diagnosis (e.g., NfL, Aβ40) track temporal location more closely, a finding also confirmed by MR. This suggests that the AD timeline may enable the identification of biomarkers of disease progression, or may itself serve as such a biomarker, as any new sample can be assigned a temporal location along that timeline. A recent study showed that p-tau217 alone can predict age of symptom onset with good accuracy^76^. Because our approach predicts age of symptom onset using temporal location on the inferred AD timeline without any AD-specific biomarkers, it may capture orthogonal information that could enhance performance when integrated with biomarker-based predictors such as p-tau217.

More broadly, accurate estimation of temporal location along the disease timeline could be transformative for therapeutic development. High failure rates in clinical trials for AD have been attributed in part to enrolling patients too late in the disease course^77^. A reliable measure of disease progression would enable direct evaluation of whether clinical trial failures are driven by late enrollment along the disease trajectory. Further, by accurately positioning individuals along the AD timeline, this framework could inform earlier and more appropriate patient selection and enable new types of clinical trial endpoints, such as preventing progression to more advanced disease states or slowing the rate of disease progression. Intriguingly, progression-related signals can be detected even in individuals without clinical symptoms. Consistent with the genetic risk model proposed above, this suggests that individuals may already be progressing along the AD timeline prior to clinical presentation, as reflected by the correct ordering of their longitudinal samples along the timeline. This supports the established hypothesis that AD disease processes begin before symptom onset^2,3^ and raises the possibility that ordering of longitudinal samples along the AD timeline could be used to identify individuals at elevated risk of developing AD symptoms.

In addition to recapitulating disease progression, our timeline inference approach enables tracking and characterization of individual trajectories over time, and quantification of how rapidly each individual progresses relative to the broader population. This is enabled by the direct comparability of the inferred timeline to the time between dates of sample collection, a feature not shared by previous methods, thereby allowing estimation of progression rates from longitudinal observations. This timeline progression rate is a machine-learning derived phenotype that characterizes the entire course of disease in an individual and is correlated with the rate of progression of clinical severity. Quantifying progression in this way enables a range of downstream applications. Prior studies have described rpAD as a distinct subtype^26,27^, with growing evidence supporting a continuum of progression rates^28,29^ that may partly reflect variation in mixed pathologies, particularly in older individuals in whom Lewy body, TDP-43, and vascular pathologies frequently co-occur with Aβ plaques and neurofibrillary tangles^78,79^. A quantitative measure for progression rate allows stratification by this progression rate on a much more granular level, allowing biological and clinical characterization across the full range of this spectrum. It also provides a basis for defining disease subtypes not by cross-sectional clinical, neuropathological, or molecular features^43,44,80^, but by the temporal dynamics of disease progression.

The importance of defining an individual-level progression rate is especially evident in the context of our proposed multidimensional paradigm of genetic disease risk. In this paradigm, progression rate is a biologically meaningful quantitative phenotype of an individual that is at least partially under separate genetic control from the potential for disease. Our approach thus extends the liability threshold model traditionally applied to case-control analyses, where susceptibility to disease is modelled as a single outcome. In this traditional model, features such as severity and progression rate are treated as secondary traits that describe differences between cases but are not informative for distinguishing cases and controls. In contrast, our results support a model where progression rate is a trait that contributes to case-control status rather than being downstream and relevant to only cases. This likely arises from the progressive nature of AD, for which a single cross-sectional measure of disease status cannot capture key features of an individual’s lifelong disease trajectory. For example, an individual with low disease potential but rapid progression may have the same overall genetic susceptibility to AD as an individual with higher disease potential but slower progression. These distinct trajectories have different biological and clinical implications, suggesting that progression is not only a modifier of disease risk but also a potential target for therapeutic intervention.

Separating disease potential from progression risks reveals distinct genetic architectures, with AD timeline progression-associated loci differing from those identified in GWAS for observed AD case-control status. Notably, we did not detect a prominent signal at the *APOE* locus, and GWAS results with and without adjustment for *APOE* genotype were highly concordant, indicating that AD timeline progression-associated variants are largely independent of this major risk factor. Previous genetic studies have only found a minor role for *APOE4* in promoting AD progression, while it has a significant effect on case-control status and age at onset^30–33^. Taken together, these findings support the notion that *APOE* primarily influences AD potential rather than the rate of AD progression.

Among the loci identified, *RDX* emerged as the most strongly supported candidate gene, with convergent evidence from gene-based association, eQTL mapping, and chromatin interaction analyses. Although it has been found to be a network hub significantly upregulated in AD^81,82^, *RDX* has not previously been implicated in AD genetics, providing an example of progression-based analyses potentially uncovering novel components of disease biology. Indeed, its *Drosophila* ortholog apparently influences locomotor behavior independent of Aβ or tau, implying contributions to neuronal maintenance during aging^83^. Furthermore, using longitudinal gene expression studies in drosophila, *RDX* was identified as a driver of a module previously associated with oligodendrogial functions in AD^83^. *RDX* encodes Radixin, a cytoskeletal protein that links actin filaments to the plasma membrane^84^. Disruption of the cytoskeleton is a well-known feature of AD pathology, most notably in the context of neurofibrillary tangles containing microtubule-associated protein tau, and actin-containing structures have also been recognized as important features to the disease^85^. Moreover, actin cytoskeleton dynamics are critically involved in microglial efferocytosis (i.e., the phagocytosis and processing of cellular debris) which is an AD pathogenic hub driven by multiple AD risk alleles and genes^86^. Several other genes prioritized by our analysis encode cytoskeletal proteins with high expression in brain, including *DOCK2* and *KIAA1279*, suggesting a possible role for these cytoskeletal structures in AD progression^87,88^. Other genes with plausible connections to known AD biology include *TLR4*, which is involved in the innate immune response to Aβ^89^, and *SEMA6D*, a neuronally expressed gene that mediates neuronal-microglia cross-talk through *TREM2* in AD and regulates microglial activation and Aβ clearance^90^. Importantly, these findings have potential therapeutic implications. While many current strategies aim to reduce the likelihood of disease, targeting pathways that influence progression rate may offer an alternative, potentially more attainable approach: slowing or stopping AD progression. Notably, several of the genes mentioned above are likely to be tractable as drug targets, and *TLR4* in particular is targeted by multiple compounds that have reached Phase III clinical trials^91^. In this context, progression-associated genes may highlight potential drug targets that act to modulate the speed of disease progression, providing opportunities to alter the clinical course of AD in ways not accessible through interventions focused on susceptibility as defined by case-control GWAS.

Taken together, our results support a multidimensional view of total AD risk in which disease arises from the interplay of at least two distinct components: risk for AD potential and risk for rate of AD progression. In this paradigm, progression rate represents a genetically influenced axis of disease biology that is largely independent of canonical susceptibility loci, yet contributes to clinical outcomes and observed case-control status. This conceptual separation of risk for potential and risk for rate of progression has implications beyond AD. Many neurodegenerative and chronic diseases are inherently progressive, and their clinical manifestations depend not only on whether pathological processes are initiated, but also on how rapidly they evolve^92–97^. A multidimensional model incorporating both potential and progression may therefore provide a more accurate representation of disease biology across a range of conditions. Variations of this framework could include nonlinear interactions between components, threshold effects, or additional axes capturing resilience or compensatory mechanisms, highlighting that total disease risk may generally be better understood as a dynamic process rather than a static state. Consistent with this broader class of models, integrating polygenic risk scores for observed case-control status and progression rate significantly improved prediction of AAO in both our data as well as in an independent larger cohort, supporting the notion that multiple genetic components contribute to disease risk, although not uniquely supporting a specific formulation of their relationship. One important implication of this framework is its possible relevance to the longstanding problem of missing heritability. One proposed explanation for missing heritability is collapsing genetically heterogeneous traits into one measurement^98^. Traditional observed case-control GWAS do exactly this, aggregating multiple biological processes into a single phenotype, which dilutes genetic signals that act specifically on disease progression or other dynamic aspects of disease. By integrating longitudinal information into genetic studies, we can decompose disease risk into multiple components to uncover genetic contributions that are obscured in conventional analyses.

## Limitations

Several limitations of this study should be considered. Our analyses were conducted in a modest sample size (1,341 individuals with genetic and longitudinal molecular data) and were restricted to individuals of predominantly European ancestry, limiting power for genetic analyses, including heritability estimation and MR. Larger and more diverse cohorts will be essential to improve power and generalizability. Our approach relies on longitudinal molecular data with multiple timepoints per individual, which remain relatively rare in the field. Power is further weakened by testing five different modalities separately, requiring a multiple test correction that would leave none of our loci genome-wide significant. Progression rate estimates also depend on sampling density and follow-up duration, may be sensitive to sparse or uneven sampling, and assume approximately linear change over the observed interval, which may not hold across all disease stages. While partially mitigated by the depth of sampling in ADNI and by extensive clinical validation of inferred AD timelines and AD timeline progression rates presented here, replication is currently limited by the scarcity of comparable longitudinal datasets. Future work with increased sampling density and longer follow-up will be important to refine these estimates and enable modeling of temporal location-dependent and nonlinear changes in AD progression rate. The performance of inferred AD timelines cannot be directly compared to those of external timelines due to the absence of fixed reference points throughout the disease or any other way of constructing a ground truth AD timeline. Although we address this through an intrinsic performance metric and multiple orthogonal validation strategies, future work incorporating additional data modalities and external benchmarks will improve interpretability and comparability. Our study primarily uses blood-based molecular data and includes limited representation of the preclinical phase, as most samples were collected after diagnosis. This is mitigated by the non-trivial number of samples collected before conversion to AD in participants enrolled before diagnosis. Extending this framework to other tissues and earlier disease stages will be important to capture the full timeline of AD. This could include autosomal dominant AD (ADAD) or patients with Down syndrome, conferring the unique benefit of knowing that individuals with these penetrant genetic variants will develop disease. Because age and disease progression are inherently colinear within individuals, age cannot be directly accounted for in AD timeline inference. Instead, we use a gcPCA framework in which CN individuals serve as a background population, enabling the isolation of disease-specific variation not present in CN. This approach effectively attenuates aging-related signals and allows capture of disease progression-specific dynamics. Finally, genetic analyses of machine learning-derived phenotypes introduce additional considerations. While GWAS signals were well-controlled and replicated across independent datasets, associated loci require independent validation and functional follow-up to confirm biological relevance. MR analyses may also be affected by violations of underlying assumptions. Seeking independent confirmation of these signals is of particular importance because of the relative scarcity of previous GWAS on progression-related features, making it more difficult to validate findings computationally. More broadly, the multidimensional model proposed here represents one of several possible formulations, and while supported by our data, it does not constitute definitive proof of the underlying causal structure.

## Conclusion

In this study, we applied a data-driven timeline inference framework to AD to reconstruct disease timelines and quantify individual-specific timeline progression rates in order to understand the genetic basis of disease progression. Using this approach, we showed that temporal location captures clinical disease severity, while timeline progression rate represents a biologically meaningful dimension of AD that is at least partially under genetic control. We further identified novel genetic loci associated with AD timeline progression rate and found that the genetic architecture of AD timeline progression rate is largely distinct from that of observed AD case-control status, including *APOE*. These findings support a multidimensional paradigm for AD risk in which total genetic risk for AD reflects the combination of risk for AD potential and risk for rate of AD progression. More broadly, they establish a framework for studying AD and other progressive disorders as dynamic, time-dependent diseases rather than a static condition from which a patient is either affected or unaffected. This perspective enables a more complete representation of disease biology and clinical heterogeneity. The identification of progression-associated genetic loci provides new insights into mechanisms that regulate how disease unfolds over time, highlighting potential therapeutic targets that may modify disease trajectory rather than disease presence. In addition, incorporating progression dynamics may improve prediction of patient outcomes, refine biomarker development by distinguishing markers of disease presence from those of disease progression, and inform clinical trial design by enabling stratification of individuals based on both disease stage and progression rate as well as by providing novel trial endpoints that measure changes in disease progression. Such distinctions may be critical for identifying patients most likely to benefit from interventions targeting different aspects of disease biology. More broadly, our results suggest that AD, and other progressive diseases, may be better understood through a multidimensional lens that integrates disease potential and temporal progression. Extending this paradigm to other neurodegenerative and chronic conditions may improve genetic discovery and provide a foundation for more precise and mechanistically informed models of disease risk.

## Data Availability

All data produced in the present study are either contained in the manuscript or will be made publicly available upon publication.

## Supplementary Figure Legends

**Supplementary Figure 1. Inferred Timelines are Stable Across Seeds and Informative Across Diagnostic Groups.**

**(A)** *Bar Plot of Estimated Performance of AD Timeline Inference Across Random Seeds.* The x-axis is the modality of longitudinal plasma metabolomics data used for timeline inference (FIA: serum metabolites measured by flow injection analysis, LCMS: plasma lipids measured by liquid chromatography-mass spectrometry, METABOLON: serum metabolites measured by metabolon platform, NMR: serum lipids and lipoproteins measured by nuclear magnetic resonance spectroscopy, and UPLC: serum metabolites measured by ultra-performance liquid chromatography mass spectrometry) and the y-axis the mean (± standard deviation) estimated performance (*τ_timeline_*) of AD timeline inference across 10 random seeds. **(B)** *Bar Plot of Estimated Performance of Null Timeline (Null τ_timeline_) in Individuals with AD.* To create Null timelines, timepoint labels were randomly shuffled within individuals. The x-axis is the modality of data used for timeline inference. Because *τ_individual_* is the upper bound of *τ_timeline_* (*τ_individual_* ≥ *τ_timeline_*), the y-axis displays *τ_individual_* as an upper bound for the estimated performance of timeline inference for Null timelines *τ_timeline_* . **(C & D)** *Heatmap of Correlations between Temporal Locations Along Modality Specific AD Timelines for Samples from Individuals with MCI **(C)** and CN Individuals **(D)**.* Axes are the data modality used to infer the AD timeline, color scale (defined in the legend) and numbers in each box are the Spearman’s correlation (*ρ*) from each pairwise comparison, and asterisks denote significance as follows *** = FDR ≤ 2.69 x 10^-12^e-12. **(E & F)** *Heatmap of F-statistics (ANOVA) for the Increase in Performance in the Association Between Cross-Validated (Methods) Temporal Locations Along All AD Timelines and Clinical Outcomes **(E)** and Biomarkers of AD **(F)** Relative to a Model Using Only Age at Sampling.* The x-axes are the diagnostic groups and the y axes the predicted clinical and biomarker outcomes. Color scale is the F statistic (defined in the legend) and numbers in each box are the F statistic (number of samples) for each comparison, and asterisks denote significance as follows *** = FDR ≤ 0.001, ** = FDR ≤ 0.01, * = FDR ≤ 0.05, ns = not significant.

**Supplementary Figure 2. AD Timeline Progression Rates are Informative Across Diagnostic Groups.**

**(A & B)** Heatmap of Correlations between Modality Specific AD Timeline Progression Rates in Individuals with MCI **(A)** and CN individuals **(B)**. Axes are the data modality used to infer the AD timeline, color scale (defined in the legend) and numbers in each box are the Spearman’s correlation (*ρ*) from each pairwise comparison, and asterisks denote significance as follows *** = FDR ≤ 0.001, ** = FDR ≤ 0.01, * = FDR ≤ 0.05, ns = not significant. **(C & D)** Heatmap of F-statistics (ANOVA) for the Increase in Performance in the Association Between Cross-Validated (Methods) AD Timeline Progression Rates and the Rate of Change of Clinical Outcomes **(C)** and Biomarkers of AD **(D)** Relative to a Model Using Only Age at Baseline. The x-axes are the diagnostic groups and the y axes the predicted clinical and biomarker outcomes. Color scale is the F statistic (defined in the legend) and numbers in each box are the F statistic (number of samples) for each comparison, and asterisks denote significance as follows *** = FDR ≤ 0.001, ** = FDR ≤ 0.01, * = FDR ≤ 0.05, ns = not significant.

**Supplementary Figure 3. AD Timeline Progression Rate GWAS Results Are Well Controlled for Inflation and Remain Stable after APOE Genotype Adjustment.**

**(A-E)** *Quantile-Quantile Plots of GWAS Results for AD Timeline Progression Rates.* The x- and y-axes are expected and observed -log_10_(p-values) respectively. The dotted line represents the expected distribution under the null hypothesis. Genomic inflation (λ) and number of individuals in the GWAS (N) are shown on each plot. Data modality used for timeline inference: FIA **(A)**, LCMS **(B)**, METABOLON **(C)**, NMR **(D)**, and UPLC **(E)**. **(F-I)** *Scatterplots of Correlation Between GWAS Results With and Without the Inclusion of APOE Genotype as Additional Covariates.* Facet titles denote whether -log_10_(p-values) or effect size (Z) of associations are used for comparison. The x- and y-axes are the -log_10_(p-values) or effect size (Z) for each variant tested for association without APOE genotypes (number of APOE2 and APOE4 alleles) as covariates (x-axis) and with APOE genotypes as covariates (y-axis). Orange dashed lines and solid lines are the identity line and a linear model fit to the data respectively. Dotted red lines representing genome-wide significance threshold (P ≤ 5 x 10^-8^) are shown in the -log_10_(p-values) plots. Correlation (Pearson r and Spearman ρ) and sample size (N) are listed in each plot. Data modality used for timeline inference: FIA **(F)**, LCMS **(G)**, METABOLON **(H)**, NMR **(I)**, and UPLC **(J)**. **(K)** *Bar Plots of Correlation Between GWAS Results With and Without Adjusting for APOE Genotype Covariates After LD-Pruning*. Plots represent either effect size (Z) or significance (-log_10_(p-values)) (facet titles). The x- and y-axes are data modality used for timeline inference and the Spearman’s correlation (ρ).

**Supplementary Figure 4. Gene-Based Association Tests for LCMS and Replication of GWAS Results Across Data Modalities.**

**(A)** *Manhattan Plot of Gene-Based Association Results for AD Timeline Progression Rate in the LCMS Modality*. The x- and y-axes are genomic locations and -log_10_(p-values) for genes tested for association with AD timeline progression rate in the LCMS modality. The dotted red line shows Bonferroni-corrected (2.6 x 10^-5^) significance. **(B)** *Quantile-Quantile Plot of Gene-Based Association Results for AD Timeline Progression Rate in the LCMS Modality.* The x- and y-axes are expected and observed -log_10_(p-values) for genes tested for association with AD timeline progression rate in the LCMS modality. The dotted line represents the expected distribution under the null hypothesis. **(C)** *Replication of GWAS Results for AD Timeline Progression Rate Across Modalities*. The x- and y-axes are the data modality of the GWAS results used as discovery (P < 5 x 10^-5^) and replication (P < 0.05) cohorts for a one-sided binomial test for replication. Color and percentage labels show replication rate and asterisks denote significance (FWER-adjusted p-value) as follows *** = P ≤ 0.001, ** = P ≤ 0.01, * = P ≤ 0.05, ns = not significant.

## Supplementary Table Descriptions

**Table S1. ADNI cohort information.**

Full Cohort, ADNI1 Cohort, ADNI2 Cohort.

**Table S2. Inferred AD timeline and AD timeline progression rate associations with cognitive metrics and biomarkers.**

Tau Timeline, Null Tau Timeline, Timeline Modality Corr, Timeline Cog Univar, Timeline Cog F-test, Timeline Bio Univar, Timeline Bio F-test, Timeline Yr Until Symptom Onset, Tau Individual Across Dx Groups, Slope Modality Corr, Timeline Slope Cog Slope Univar, Slope Cog F-test, Timeline Slope Bio Slope Univar, Slope Bio F-test, Timeline vs Age Correlation.

**Table S3. GWAS summary statistics filtered at 1e-3.**

GRCh38_FIA_without_APOE, GRCh38_UPLC_without_APOE, GRCh38_NMR_without_APOE, GRCh38_LCMS_without_APOE, GRCh38_METABOLON_without_APOE, GRCh38_FIA_with_APOE, GRCh38_UPLC_with_APOE, GRCh38_NMR_with_APOE, GRCh38_LCMS_with_APOE, GRCh38_METABOLON_with_APOE, GRCh37_FIA_without_APOE, GRCh37_UPLC_without_APOE, GRCh37_NMR_without_APOE, GRCh37_LCMS_without_APOE, GRCh37_METABOLON_without_APOE, GRCh37_FIA_with_APOE, GRCh37_UPLC_with_APOE, GRCh37_NMR_with_APOE, GRCh37_LCMS_with_APOE, GRCh37_METABOLON_with_APOE.

**Table S4. Mendelian randomization and other post-GWAS analyses.**

QQ_Stats, APOE_Scatter_Stats, APOE_LD_Pruned_Correlation, External_Sumstats_Info, External_Replication, Cross_Modality_Replication, MR_Forward_1Sample, Age_of_Dx_Model.

**Table S5. FUMA input parameters and full gene mapping and fine-mapping results**

FUMA_Params_FIA, FUMA_Params_LCMS, FUMA_Params_METABOLON, FUMA_Params_NMR, FUMA_Params_UPLC, Full_Gene_Results, Fine_Mapping_Results.

## Methods

### ADNI Dataset

Data used in the preparation of this article were obtained from the Alzheimer’s Disease Neuroimaging Initiative (ADNI) database (adni.loni.usc.edu). The ADNI was launched in 2003 as a public-private partnership, led by Principal Investigator Michael W. Weiner, MD. The primary goal of ADNI has been to test whether serial magnetic resonance imaging (MRI), positron emission tomography (PET), other biological markers, and clinical and neuropsychological assessment can be combined to measure the progression of mild cognitive impairment (MCI) and early Alzheimer’s disease (AD). For up-to-date information, see https://www.adni-info.org.

ADNI classifies subjects into diagnostic groups based primarily on the results of cognitive and clinical tests such as Mini-Mental State Examination scores (MMSE; AD: 20 - 26, MCI and CN: 24 - 30) and Clinical Dementia Rating (CDR; AD: 0.5 - 1.0, MCI: 0.5, CN: 0)^35^. To be classified as MCI or AD, subjects must have a valid memory complaint^35^. To be classified as AD, subjects must meet the National Institute of Neurological and Communicative Diseases and Stroke / Alzheimer’s Disease and Related Disorders Association (NINCDS/ADRDA) criteria for probable AD^35,99^. Additional criteria for enrollment and diagnostic classification can be found on the ADNI website (adni.loni.usc.edu).

The five following sparse longitudinal blood metabolomics data from ADNI were used: 1) serum metabolites measured by flow injection analysis (FIA), 2) serum metabolites measured by ultra-performance liquid chromatography mass spectrometry (UPLC), 3) serum lipids and lipoproteins measured by nuclear magnetic resonance spectroscopy (NMR), 4) plasma lipids measured by liquid chromatography-mass spectrometry (LCMS), and 5) serum metabolites measured using the metabolon platform (METABOLON). For each individual, multiple longitudinal samples were collected, with measurements from these five modalities generated for largely overlapping sets of individuals and, in most cases, from blood samples obtained at matched timepoints. For each individual, all samples were assigned the same diagnostic group based on the individual’s clinical diagnosis from the “adnimerge” object in the ADNIMERGE R package (version 0.0.1)^100^ according to the following hierarchical assignment: samples from individuals with at least one instance of dementia diagnosis were classified as AD; samples from the remaining individuals with at least one instance of MCI diagnosis were classified as MCI; and the remaining samples were classified as cognitively normal (CN). The five following clinical assessments were also obtained for each individual at multiple timepoints: 1) Clinical Dementia Rating Sum of Boxes (CDR-SB), 2,3) Alzheimer’s Disease Assessment Scale-Cognitive Subscale 11-item and 13-item versions (ADAS11, ADAS13), 4) Mini-Mental State Examination (MMSE), and 5) Functional Activities Questionnaire (FAQ). These clinical assessments were obtained from the “adnimerge” object in the ADNIMERGE R package (version 0.0.1)^100^ and many clinical assessments were obtained from matching timepoints as the longitudinal blood metabolomics data. The five following sparse longitudinal blood plasma biomarkers from ADNI were used: 1) p-tau217, 2) Aβ42, 3) Aβ40, 4) Aβ42/Aβ40 ratio, and 5) NfL. These plasma biomarkers were obtained from timepoints which matched some of the timepoints from the longitudinal blood metabolomics data. Genotype data were obtained from ADNI for two cohorts: ADNI1^101^ and ADNI2/GO. Briefly, ADNI1^101^ comprised 662 individuals genotyped using the Illumina Human610-Quad BeadChip with genotype data aligned to hg18 and ADNI2 comprised 680 individuals genotyped using the Illumina HumanOmniExpress BeadChip with genotype data aligned to hg19. All corresponding LONI-IDA file names and dates of download for ADNI data used in the preparation of this manuscript are listed in the “*Data and Code Availability*” section.

### Preprocessing of Longitudinal Metabolomics Data

Preprocessing for serum metabolites data modalities measured by FIA and UPLC was performed using a custom adaptation of the approach reported for the corresponding non-longitudinal data modalities^102^. Metabolite columns with >40% missingness were removed. Non-zero values below the limit of detection (LOD) were set to half of the plate-specific LOD for each metabolite from as defined in the FIA and UPLC quality control data sheets (LONI-IDA file name and date of download are listed in the “*Data and Code Availability*” section). Since missing and zero values in serum metabolite data often reflect the absence of a measure rather than the absence of the metabolite^103,104^, each metabolite with missing or zero were imputed as the mean of non-missing, non-zero values for that metabolite^105^. All metabolite features were log2-transformed. All metabolite features were batch corrected using a previously published approach which fits a linear mixed model to the data with random effects for sample ID and plate ID; then subtracts out the component of variance attributed to plate ID^106^. Standard pooled quality control (SPQC) samples from the FIA and UPLC quality control data sheets (LONI-IDA file name and date of download are listed in the “*Data and Code Availability*” section) served as technical replicates across plates.

The serum metabolites as measured by METABOLON contains metabolites measured by four different platforms, each with its own batch information, and were thus processed using the same approach as the one described for FIA and UPLC with the following exceptions: 1) batch correction was performed separately for each platform, with Platform ID identified from the “TEXT” column of the ADNI data dictionary (LONI-IDA file name and date of download are listed in the “*Data and Code Availability*” section); and 2) All non-zero values were not modified for LOD because LOD information was not available.

The serum lipids and lipoprotein as measured by NMR were processed using the same approach as the one described for FIA and UPLC with the following exceptions: 1) Since LOD information was not available, zero and missing values were imputed as half of the minimum non-zero, non-missing value, a common approach for lipidomics data processing^107^; 2) No batch correction was performed because technical replicates across plates were not available. Since this modality included many derived features such as ratios calculated from multiple measured features, Hartigan’s dip test was used to test for unimodal distribution using the “dip.test” function from the “diptest” (v0.77.1)^108,109^ R package. Any features which were not unimodal and log-unimodal were removed from downstream analyses.

The plasma lipidomics as measured by LCMS had already undergone certain preprocessing steps such as batch alignment (corresponding methodological information was obtained from the “ADMC Lipidomics Meikle Lab Longitudinal Data Matrix Methods [ADNI1,GO,2] (PDF)” file in LONI-IDA on May 21, 2026)^110^. Additionally, no technical replicates across batches were available, no LOD information was available, and no missing or zero values were present. Thus, no steps from the approach described for FIA and UPLC were conducted.

After preprocessing, all modalities were munged and formatted as follows. For any individuals with multiple data points available for samples collected at the same exam date, the first occurring data point was kept ensuring there was only one data point per exam date per individual. Individuals without longitudinal data (i.e., individuals with less than two unique samples) were removed.

### Timeline Inference Method

To infer AD timelines, we applied our previously developed Timeline Inference Method (TIM) framework^34^. TIM takes as input features measured longitudinally across multiple individuals along with individual identifiers and time since first sampling for each sample; and outputs an estimated timeline consisting of temporal locations for each sample aligned along a single axis across all individuals. TIM also estimates the Kendall’s *τ* correlation between our inferred timeline and the true unknown timeline and reports this statistic, 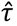_timeline_, which ranges from 0 (random ordering of samples in relation to true ordering) to 1 (perfect reproduction of the true ordering of samples)^34^.

Briefly, the TIM framework uses a cross-validated machine learning approach to fit an elastic net regression model with regularized coefficients for each input feature and non-regularized intercepts for each individual^34^. Model hyperparameters are optimized via nested cross-validation using a custom non-parametric statistic we previously developed^34^. This statistic, 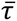_individual_, quantifies how well the inferred ordering reproduces the known true ordering of samples within each subject, regardless of the time intervals between samples. This statistic is calculated as the Kendall’s *τ* correlation between the temporal location along the inferred timeline and the time since first sampling within individuals, as a weighted mean across all individuals with each individual’s weight equal to the number of sample pairs for that individual.

Across all five data modalities, model training and evaluation was conducted using AD individuals only. After model training was complete, all samples from subjects with AD, MCI, and CN were then projected onto the inferred timeline of AD using the trained model. All timelines used a single random seed (seed = 1).

### Generalized Contrastive Principal Component Analysis

To account for the variation in the longitudinal metabolomics data attributable to aging, generalized contrastive principal component analysis (gcPCA)^36,37^ was performed prior to running TIM. Data generated from samples from individuals with AD were defined as the target dataset and data generated from samples from CN individuals were defined as the background dataset for gcPCA. Generalized contrastive principal components (gcPCs) were used as input features for timeline inference.

### Calculating Individual-Specific AD Timeline Progression Rates

Using an approach we previously developed, AD timeline progression rate was calculated for each individual as the slope of the linear regression between inferred temporal location along AD timelines and years since first sample for the samples from that individual^34^. This slope represents the AD timeline progression rate as inferred timeline units per calendar year. AD timeline progression rates were computed separately for each data modality and diagnostic group.

### Evaluating Timeline Stability and Estimating Null Distributions

Stability of estimated performance for inferred AD timelines across fold splits was estimated by repeating timeline inference procedure across 10 random seeds (seeds = 1-10) for each data modality. Null distributions for 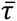_individual_ were estimated in each data modality using the following procedure: 1) timepoint labels were randomly shuffled within individuals; 2) a null AD timeline was inferred using the shuffled timepoint labels and 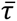_individual_ estimated^34^; 3) this procedure was repeated for 101 random seeds (seeds = 1-101) for random shuffling while using the same random seed (seed = 1) for fold splitting for timeline inference. Since 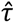_timeline_ ≤ 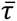_individual_ by definition, this distribution is a valid (and more computationally efficient) upper bound for the null distribution for 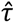_timeline_34.

### Cross-Validation Framework to Generate Out-Of-Sample Timeline Metrics

Holdout sets for testing associations between inferred timelines and features of interest (within individual sample ordering, clinical metrics, biomarkers, and years until symptom onset) were created using the following procedure. AD individuals were partitioned into three holdout folds of approximately equal numbers of individuals. For each of holdout fold, a timeline of AD was inferred using the data from the remaining individuals with AD with the full CN set as the background for gcPCA. Individuals in the holdout set were then projected onto this inferred timeline, yielding out-of-sample temporal locations along these timelines for metabolomics samples from these individuals. This process was repeated for all three holdout folds, thereby yielding out-of-sample temporal locations on the AD timeline for all samples from AD individuals. Samples from MCI and CN individuals were projected onto each of the three timelines inferred from each fold and then averaged together to obtain a single temporal location along the AD timeline for each sample from MCI and CN individuals. This process was repeated across 10 random sampling seeds (seeds = 1-10) for fold partitioning to ensure stability of results. For each random sampling seed, timeline inference was performed with the same seed. AD timeline progression rates were calculated across all 10 seeds and then averaged together to obtain one AD timeline progression rate per individual for association testing. Additionally, timeline locations were averaged together across all 10 seeds to obtain one timeline location per sample for association testing.

### Assessing Within-Individual Sample Ordering

To test whether inferred timelines preserve the known within-individual sample order significantly above chance, we used our previously developed statistical test (block *τ* test)^34^. This test calculates the 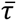_individual_ and evaluates it against the null hypothesis of random within-individual sample order in relation to true time. This test was applied separately for each data modality and diagnostic group (AD, MCI, CN), using the out-of-sample timeline predictions from each of the 10 random sampling seeds described in the “*Cross-Validation Framework to Generate Out-Of-Sample Timeline Metrics*” section of the methods.

### Association Testing for Clinical and Biomarker Features

Inferred temporal locations along the AD timelines as described in the “*Cross-Validation Framework to Generate Out-Of-Sample Timeline Metrics*” section of the methods were tested for association with clinical severity (CDR-SB, ADAS11, ADAS13, MMSE, FAQ) and plasma biomarkers of AD (p-tau217, Aβ42, Aβ40, Aβ42/Aβ40, NFL) for each modality of data (FIA, LCMS, NMR, UPLC, METABOLON) and diagnostic group (AD, MCI, CN) using linear models while accounting for age at sampling. The following formula was used:

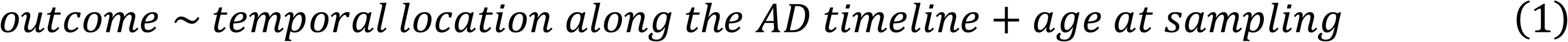

For each clinical metric of severity and plasma biomarker of AD, rate of change per year was calculated for each individual with at least two measures for each respective feature over time using the same approach as the ones described to calculate AD timeline progression rates (see section: “*Calculating Individual Rates of AD Progression*”). Associations between these rate of progression outcomes and AD timeline progression rates as described in the “*Cross-Validation Framework to Generate Out-Of-Sample Timeline Metrics*” section of the method were tested using a linear model while accounting for age at baseline. The following formula was used:

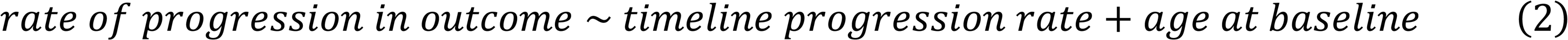

### Association Testing between Temporal Location along AD Timeline and Time Until Symptom Onset

Time until symptom onset was calculated as previously described^76^. Briefly, symptom onset was defined as the first age of reported non-zero CDR-SB for any individual with a CDR-SB score of zero at study enrollment. Years until symptom onset for each visit was calculated as the difference between age of symptom onset and age at visit. A multivariate linear model was fit for years until symptom onset using temporal location along the AD timelines for all data modalities as described in the “*Cross-Validation Framework to Generate Out-Of-Sample Timeline Metrics*” section of the method as predictors. The following formula was used:

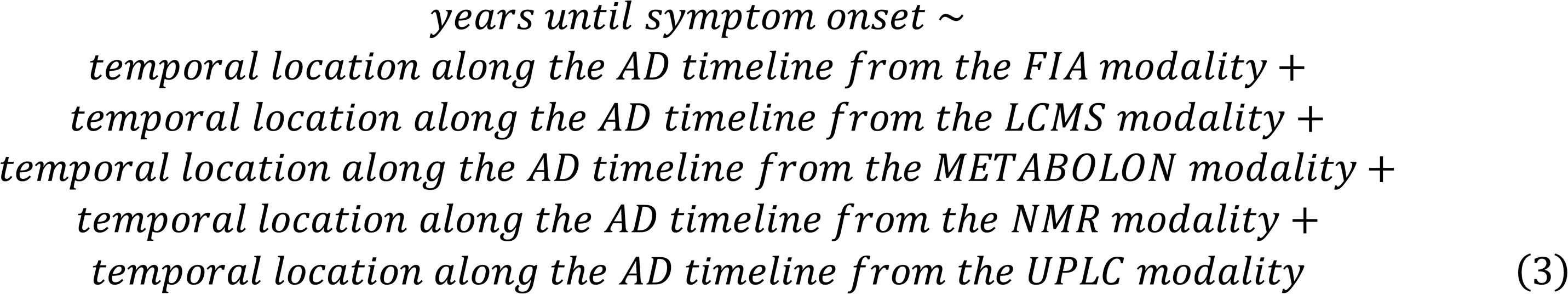

### Association Testing across All Modality-Specific AD Timelines

To assess whether temporal location along AD timelines from all five metabolomics data modalities as described in the “*Cross-Validation Framework to Generate Out-Of-Sample Timeline Metrics*” section of the method jointly predicted clinical and biomarker outcomes significantly better than age at sampling alone, F-tests were performed using the R “anova()” function. There, a full model containing cross-validated temporal location along AD timelines from each modality and age at sampling as predictors was compared against a null model containing only age at sampling as a predictor. This approach was also used to assess whether AD timeline progression rates from all five metabolomics data modalities as described in the “*Cross-Validation Framework to Generate Out-Of-Sample Timeline Metrics*” section of the method jointly predicted rate of progression outcomes as defined in the section “*Association Testing for Clinical and Biomarker Features*” of the method significantly better than age at baseline enrollment alone.

### Genotype and Covariate Data Used for Genetic Association Testing

Raw genotype data from the ADNI1 cohort (hg18) and the ADNI2/GO cohort (hg19) were lifted over to hg38 using the UCSC LiftOver (v24-Jan-2025) tool^111–113^, with unmapped variants removed.

The covariate matrix for association testing consisted of the following variables for each individual: 1) sex, 2) age at baseline, 3) age at baseline * sex, 4) and the time interval elapsed between the first and last sample collection visit dates.

### Genotype Imputation

Prior to imputation, hg38 genotype files were harmonized to the reference genome using the bcftools (v1.22)^114^ “+fixref” plugin with the mode (“-m”) set as “top”. They were then split by chromosome using bcftools (v1.22)^114^. Genotype imputation was conducted using Michigan Imputation Server 2^115^ on September 22, 2025 with the following parameters: 1) Reference Panel set to “1000G Phase 3 (GRCh38/hg38) [BETA]”^116–119^, 2) Array Build set to “GRCh38/hg38”, 3) rsq Filter set to “off”, 4) Phasing Engine set to “Eaglev2.4 (phased output)”^120,121^, 5) Allele Frequency Check set to “Off”, 6) and Mode set to “Quality Control & Imputation”. Imputed dosages from Michigan Imputation Server 2 were converted from Variant Call Format (VCF) to PLINK binary format (.bed, .bim, .fam) using PLINK (v1.90b6.21)^122–124^ with the “--keep-allele-order” flag to preserve major and minor allele definitions from the imputation server.

### Genome-Wide Association Testing

Individual-specific AD timeline progression rates for each of the 5 metabolomics data modalities estimated with seed 1 were used as continuous traits tested for association with genetic variation. For each data modality, a separate GWAS were performed for six subcohorts of ADNI, consisting of the unique combination of genetic cohorts (ADNI1, ADNI2/GO) and diagnostic groups (AD, MCI, CN). AD timeline progression rates for each subcohort were separately transformed using an inverse rank normalized transformation (IRNT). REGENIE (v3.2.2)^125^ was used for association testing. For REGENIE step 1, the following additional genotype processing was conducted separately for each unique combination of subcohort and modality with PLINK (v1.90b6.21)^122–124,126,127^: 1) individuals without phenotypes were removed; 2) variants with a minor allele frequency (MAF) < 1% were removed; 3) variants with a Hardy-Weinberg equilibrium exact test p-value < 1e^-6^ were removed^126,127^; 4) variants with missingness > 10% were removed; 5) samples with missingness > 10% were removed; 6) LD pruning was conducted with a window size of 1000, a step size of 100, and an r^2^ threshold of 0.9; 7) three specific variants were excluded due to perfect collinearity with covariates: rs12734338 (collinear with Sex), rs7412 (collinear with APOE2), and rs3883013 (also collinear with Sex^128^); 8) and variants not on autosomes or chromosome X were removed. PCA was conducted on the resulting genotypes using PLINK (v1.90b6.21)^122–124^ and the top 10 PCs are added to the covariate matrix for both steps of REGENIE^125^. Step 1 and 2 of REGENIE were conducted using REGENIE (v3.2.2)^125^ with a respective block size of 1000 and 200. Modality-specific results were meta-analyzed across subcohorts using METAL (v2018-08-28)^129^ using default settings (“AVERAGEFREQ” and “TRACKPOSITIONS” were set as “ON” to ensure allele frequency, chromosome, and base pair information were present in the meta-analyzed output).

### Summary Statistics Munging

Summary statistics munging was performed using the “format_sumstats” function from the MungeSumstats (v1.19.6, commit 5dfb8a62cabc8de8596f4952e341f9bcbe10065e)^130,131^ R (v4.4.1)^132^ package with the “ref_genome” argument set to “GRCh38”. Reference genomes were loaded using the “SNPlocs.Hsapiens.dbSNP155.GRCh38” (v0.99.24)^133^ and “BSgenome.Hsapiens.NCBI.GRCh38” (v1.3.1000) R (v4.4.1)^132,134^ packages. The following changes were made from the default arguments in the “format_sumstats” function: 1) the “bi_allelic_filter” argument was set to FALSE to avoid unnecessary variant filtering, and the “flip_frq_as_biallelic” argument was set to TRUE to allow proper allele-frequency flipping during harmonization; 2) to preserve alleles for downstream analysis, the “on_ref_genome” and “allele_flip_drop” arguments were set to FALSE; and 3) “N_std” argument was set to “Inf” to disable N-based filtering of alleles.

After the “format_sumstats” function, any variant which had only been present in one cohort before meta-analysis was identified and removed. Whenever absent, standard error (SE) and effect size (BETA) values were calculated from Z-score (Z), sample size (N), allele frequency (FRQ), using the following equations:

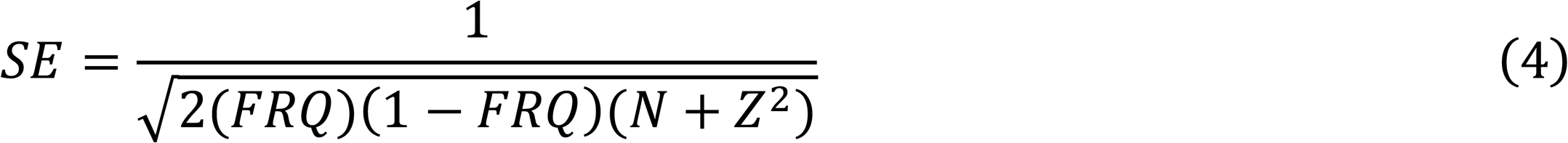

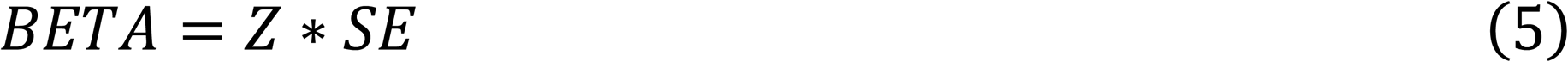

### Functional Annotation and Gene Mapping (FUMA)

Functional annotations, positional gene mapping, expression quantitative trait locus (eQTL) gene mapping, three-dimensional (3D) chromatin interaction gene mapping, and gene-based association testing was conducted using the Functional Mapping and Annotation of Genome-Wide Association Studies (FUMA) (v.1.8.3) platform and the tools contained within including Multi-marker Analysis of GenoMic Annotation (MAGMA) (v.1.10) and Annotate Variation (ANNOVAR) (v.2017-07-17)^57,58,135^.

Summary statistics were lifted over into GRCh37 prior to uploading to FUMA. Liftover was conducted using the “liftover” function from the MungeSumstats (v1.19.6, commit 5dfb8a62cabc8de8596f4952e341f9bcbe10065e)^130,131^ R (v4.4.1)^132^ package with the “ref_genome” argument set to “GRCh38” and the “convert_ref_genome” set to “GRCh37”. Reference genomes were loaded using the “SNPlocs.Hsapiens.dbSNP155.GRCh38” (v0.99.24)^133^, “BSgenome.Hsapiens.NCBI.GRCh38” (v1.3.1000)^134^, and “SNPlocs.Hsapiens.dbSNP155.GRCh37” (v0.99.24)^136^ R (v4.4.1)^132^ packages. All FUMA input parameters are listed in **Supplementary Table 5**.

### Gene Mapping and Fine-mapping

To identify the lead variants with genome-wide and suggestive associations (**Table 1**) from the munged GRCh37 summary statistics (the same summary statistics which were uploaded to FUMA), a distance-based LD-clumping procedure was applied to all loci with P < 1e-5 as follows: 1) any SNPs within 500kb of the most significant SNP were removed, 2) this process was repeated iteratively for each SNP starting from the most significant SNP to the least significant one according to their p-value. Lead SNPs which did not have supporting LD evidence (at least one SNP within 500kb and P < 1e-4) were removed. This analysis was conducted separately for each GWAS. All lead SNPs from each GWAS are shown in **Table 1**.

To identify one prioritized gene per locus, each suggestive lead variant was mapped to a candidate gene as follows. Genes with a higher number of types of evidence from the following evidence type categories were prioritized: 1) at least one SNP was positionally mapped within 10kb to that gene, 2) at least one SNP was an eQTL for that gene, 3) at least one SNP was mapped to that gene via chromatin interaction analysis, and 4) that gene has a MAGMA gene-based-p-value of less than 0.05. If multiple candidate genes in a given locus had an equal number of evidence types, then genes with a “protein_coding” class were prioritized. In the scenario where there are still multiple genes per locus, then genes with the lowest eQTL FDR and then genes with the lowest MAGMA gene-based-p value were prioritized. Finally, we considered the minimum GWAS P-value among mapped SNPs (from the “minGwasP” column) to prioritize and distinguish amongst any remaining occurrences of multiple candidate genes. For any loci which had no valid gene to prioritize, cytobands from the UCSC genome browser^111–113^ were used in place of gene labels.

Fine-mapping of GRCh37 lead SNPs (**Table 1**) was conducted using the Statistical And Functional Fine-mApping of GWAS Risk locI (SAFFARI)^56^ pipeline to run the following four fine-mapping methods: 1) SuSiE^137^, 2) FINEMAP^138^, 3) PolyFun^139^ + SuSiE^137^, and 4) PolyFun^139^ + FINEMAP^138^. Functional priors for PolyFun were estimated using the precomputed baseline-LF 2.2.UKB model^139^. The maximum number of causal SNPs was set to 5 with “--max-num-causal” flag and precomputed UK Biobank LD matrices were specified with the “--ld” flag^140^. Fine-mapping results were subsequently processed to maintain only fine-mapped SNPs with posterior inclusion probability (PIP) of greater than 50% and were within a 95% credible set. For these fine-mapped variants, gene mapping was conducted using ENSEMBL Variant Effect Predictor (VEP) (v112)^141^, and maintaining those convergently identified by at least two of the four fine-mapping methods that SAFFARI runs (**Table 1**).

### Estimating Genetic Heritability and Computing Genetic Correlation

Multiple attempts were made to estimate genetic heritability and compute genetic correlation between our GWASs of AD timeline progression rate and existing GWASs in the GWAS catalog. These attempts included the LD Score Regression (v1.0.0) “ldsc.py” script with the “--rg” command^49,142^, the LDAK (v5.2)^50,143,144^ software with the “--sum-cors” argument, and the online power calculator (http://spark.rstudio.com/ctgg/gctaPower) for GCTA-GREML^51,54^.

### Replication in External GWAS

To evaluate whether our findings replicated in external GWAS, we used summary statistics from the GWAS catalog^145^. For all studies used, the GWAS catalog accession ID and date of download are listed in the “*Data and Code Availability*” section; and links to the corresponding papers for each GWAS catalog accession ID are listed in **Supplementary Table 4**. Summary statistics for each GWAS catalog study were downloaded from the location listed in **Supplementary Table 4** and were munged as described in the “*Summary Statistics Munging*” section.

For each modality, SNPs with P < 5 x 10^-5^ were identified and selected as candidate SNPs for replication. SNPs were considered replicated in an external study if they both reached nominal significance (P < 0.05) and showed a concordant direction of effect. For external traits expected to decrease with an increased rate of AD progression (i.e. external phenotypes with opposite directions of effect than our phenotype), the direction of effect was flipped prior to replication analysis.

A one-sided binomial test was used to determine whether the observed number of replicated SNPs was greater than what would be expected under the null hypothesis (probability of 0.025). The “pbinom()” R function with the “lower.tail” argument set as “FALSE” was used for the binomial test. P-values were adjusted for multiple testing (across modalities and external studies) using the Bonferroni correction to control the Family-Wise Error Rate (FWER) using the “p.adjust()” R function. Tests with no overlapping SNPs between the discovery and external GWASs, or where no discovery SNPs in the overlapping set reached the suggestive threshold (P < 5 x 10^-5^) were excluded. This framework was also applied to test replication for all possible pairwise comparisons between the AD timeline progression rate GWAS results from the five different data modalities.

### APOE Independence Analysis

To assess whether genetic associations with AD timeline progression rate were driven by the *APOE* locus, all GWAS analyses were repeated with number of APOE2 alleles and number of APOE4 alleles included as covariates. Correlation between the original GWAS results and the APOE genotype-adjusted GWAS results for each modality was calculated for both effect size (Z-score) and significance (-log10(P)). These correlations were also calculated for LD-pruned summary statistics using PLINK (v1.90b6.21)^122–124^ for LD pruning with a window size of 100, a step size of 50, and an r^2^ threshold of 0.05.

### Polygenic Risk Scores

Polygenic risk scores (PRS) for AD timeline progression rate were computed using PRSice-2 (v.2.2.6)^146^ with 1000 Genomes Phase 3^116–119^ as an external LD reference panel. PRS models included the same covariates described in the “*Genotype and Covariate Data Used for Genetic Association Testing*” section. Loci with duplicate chr:pos identifiers were excluded prior to PRS calculation.

### Mendelian Randomization

We constructed genetic instruments for MR using PRSice-2 (v.2.2.6)^146^. Parameters for PRSice were as described in the “*Polygenic Risk Scores*” section, except with a fixed p-value threshold of 5.005 × 10^-5^. One-sample MR was conducted using 2-step least squares (2SLS) regression in the ivreg (v.0.6.7)^147^ R package. In all regressions we controlled for age, sex, age-sex interaction, and time between first and last sample; in regression on the genetic instruments, we additionally controlled for genotyping cohort (ADNI1 or ADNI2) and 10 genetic principal components. The exposure variable was AD timeline progression rate, as described for univariate associations above. Outcome variables were also calculated as described for univariate associations above. Due to the lack of previously published GWAS for tested outcome variables (i.e. rates of change over time in clinical severity measures and plasma biomarkers) we were unable to conduct either MR with AD timeline progression rate as the outcome or 2-sample MR.

### Additional Formulation for Multidimensional Genetic Model of Risk

In traditional case-control GWAS of AD, risk loci are modelled as contributing to genetic liability of disease, which in turn controls susceptibility to disease:

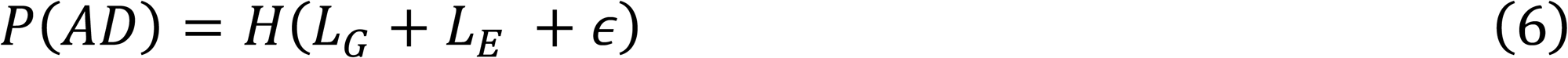

where *P*(*AD*) is probability of AD diagnosis, *L_G_* and *L_E_* are the genetic and environmental components of disease liability or susceptibility, *ε* is an error term, and *H* is the Heaviside step function, which is 0 below a specified threshold and 1 above it. Under the assumption that *L_E_* and *ε* are both normally distributed in the population, we can remove these terms and focus solely on the genetic component:

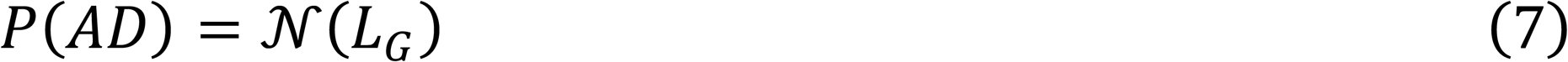

where N is a normal distribution^63–65^. Under this model, all genetic loci contribute additively to this single *L_G_* term, which in turn determines the individual’s risk of disease.

Our model replaces the univariate normal distribution operating on a single genetic liability term with a multivariate distribution combining at least two distinct components. We define these components as the risk for AD potential Φ, representing an individual’s vulnerability to the pathophysiological processes underlying AD, and the risk for rate of AD progression Δ, representing vulnerability to processes that modulate the rate at which an individual progresses along the true timeline of the disease (**Figure 6A**). This model can be expressed in a general form as:

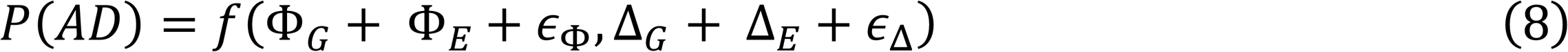

where Φ*_G_* and Φ*_E_* are the genetic and environmental components of risk for potential, Δ*_G_* and Δ*_E_* are the genetic and environmental components of risk for rate of progression, *ε*_Φ_ and *ε*_Δ_ are separate error terms for the two components, and *f* is an unspecified bivariate probability distribution. This distribution could take many forms, but one convenient possible formulation is:

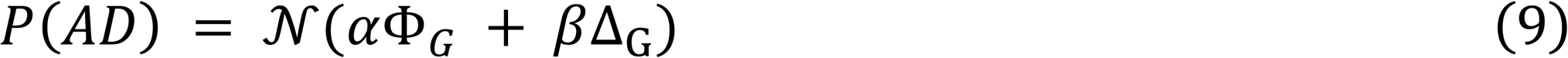

where *α* and *β* are constant coefficients defining the relative contributions of risk for AD potential and risk for rate of AD progression to overall genetic liability of AD. This formulation is identical to equation 7 but with the genetic component of disease susceptibility defined as a linear combination of the genetic components of potential and progression:

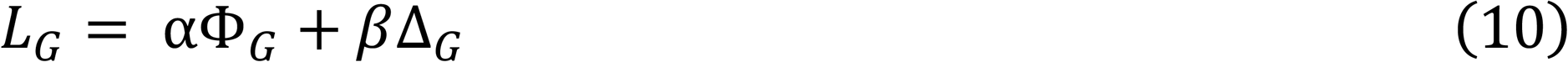

In this model, an individual may have high susceptibility to AD due to having high potential for disease (even with a relatively low progression rate), but an individual with high progression rate can also have the same high susceptibility even if potential is low. Critically, while high AD potential with slow progression and lower AD potential with rapid progression may result in similar observed case-control status, they are not equivalent: they represent distinct underlying biology, distinct strategies for disease management, and distinct therapeutic opportunities. This is only one of many possible models that combine a risk factor for AD potential with a risk factor for rate of AD progression, and different models may have different implications.

To investigate whether this model was well represented in our data, we assessed whether combining PRS from a previously published case-control GWAS^13^ and from our GWASs of AD timeline progression rate significantly improved age of diagnosis prediction compared to case-control PRS alone in the ADNI1 cohort for individuals who were enrolled as cognitively normal controls at baseline but were diagnosed with dementia over the course of the study. A likelihood ratio test was performed by computing the log likelihood and degrees of freedom for the full and null models specified by the formulas below using the “logLik()” R function and p-values were computed using the “pchisq()” R function with “q” set to twice the difference in log likelihoods between the two models, “df” equal to the difference in degrees of freedom between the two models, and “lower.tail” set to FALSE^148^.

Full model:

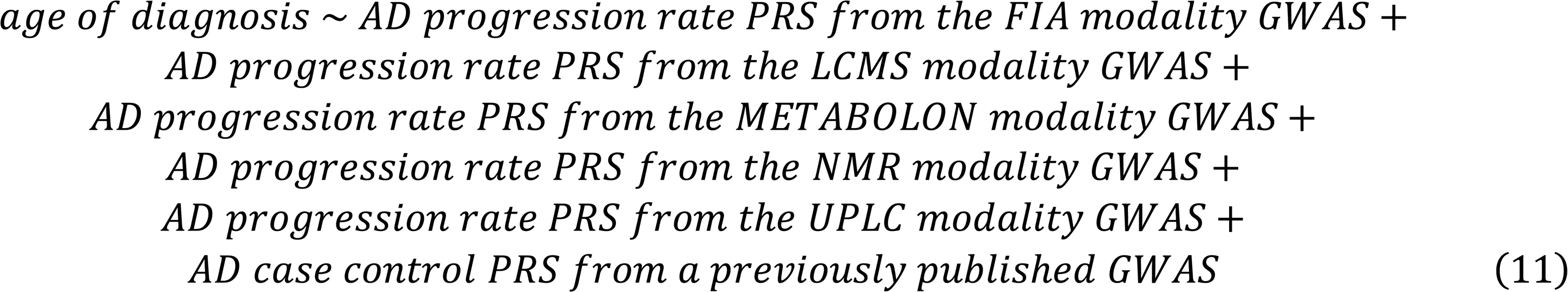

Null model:

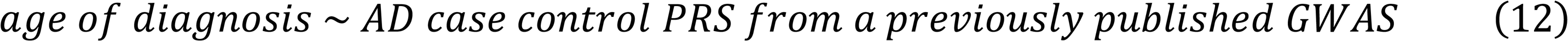

To ensure the ADNI1 cohort was not present in the training data, AD timeline progression rate PRS was computed using PRSice-2 (v.2.2.6)^146^ as described in the “*Polygenic Risk Scores*” section, except for the following adjustments: 1) GWAS results from only the ADNI2/GO cohort were used as the base summary statistics for PRS models (REGENIE results from all three disease groups within ADNI2/GO were meta-analyzed together as described in the “*Genome-Wide Association Testing*” section and munged as described in the “*Summary Statistics Munging*” section), 2) p-value permutation was conducted in the ADNI2/GO cohort with “--perm” set to “1000”. AD case control PRS was computed using PRSice-2 (v.2.2.6)^146^ as described in the “*Polygenic Risk Scores*” section, except for the following adjustments: 1) summary statistics from Bellenguez et al. (GCST90027158)^13^ were used as the base summary statistics for PRS models, 2) p-value permutation was conducted in the ADNI2/GO cohort with “--perm” set to “1000”.

### ADSP whole-genome sequencing data and study participants

Whole-genome sequencing (WGS) data were obtained from the Alzheimer’s Disease Sequencing Project (ADSP)^149^ through the National Institute on Aging Genetics of Alzheimer’s Disease Data Storage Site (NIAGADS). All participants in ADSP provided informed consent, and contributing studies received approval from their local institutional review boards. All analyses were conducted under approved NIAGADS data access agreements. Sequencing data were generated across multiple ADSP phases and harmonized through centralized joint-calling and quality-control pipelines as previously described^150^. Initial sequencing processing, alignment, variant calling, and harmonization were performed by the Genome Center for Alzheimer’s Disease (GCAD)^150^.

Analyses were restricted to individuals with available WGS data, phenotype information, and required covariates. Available phenotypic and covariate data included clinical diagnosis status, age at symptom onset, sex, self-reported race and ethnicity, APOE genotype, genetic ancestry estimates, and sequencing metadata. Individuals from the Alzheimer’s Disease Neuroimaging Initiative (ADNI) were excluded from the present analyses. Global genetic ancestry was inferred using ADMIXTURE-derived ancestry proportions^151^. Each participant was assigned to a genetic ancestry super-population according to the predominant ancestry component, defined as the genetic ancestry group with the largest estimated ancestry proportion. All analyses were performed in the full multi-ancestry cohort. For downstream association analyses of AAO, analyses were restricted to AD patients with AAO greater than 65 years.

### Sequence processing and variant quality control

ADSP WGS variant call format (VCF) files were used as the starting point for all downstream analyses. Prior to downstream analyses, additional study-specific quality-control procedures were applied. Multi-allelic variants were decomposed into biallelic representations, and variants failing quality-control criteria were removed. Genomic Data Structure (GDS) files were converted to PLINK format using seqGDS2BED from the SeqArray package in R^152,153^. Variant identifiers were harmonized to chromosome-position-reference-alternate (CPRA) format to ensure consistent variant representation across genotype data, summary statistics, and linkage disequilibrium reference panels. SNP identifiers in external summary statistics and linkage disequilibrium reference datasets were similarly converted or matched to CPRA identifiers before downstream analyses.

### ADSP Polygenic risk score analyses

Polygenic risk score analyses were performed using PRSice-2 (v.2.2.6)^146^. Summary statistics used for PRS calculation were harmonized prior to score construction. Variant identifiers were standardized to chromosome-position-reference-alternate (CPRA) format, and variants with either allele longer than one nucleotide were excluded by removing variants for which A1 or A2 exceeded 1 nucleotide. Sites with non-unique variant identifiers in the summary statistics used for PRS calculation were also removed.

External phenotype and covariate files were supplied to PRSice, including sex, genetic ancestry principal components, sequencing center or platform, diagnosis status, and AAO where appropriate. PRS analyses were conducted in the full ADSP cohort after excluding all ADNI samples. PRSice p-value inclusion thresholds were selected based on optimization in ADNI and then applied to the ADSP analyses. The optimized thresholds were 1 for FIA timeline progression rate summary statistics, 0.0195501 for LCMS timeline progression rate summary statistics, 1 for METABOLON timeline progression rate summary statistics, 0.0620001 for NMR timeline progression rate summary statistics, 0.339 for UPLC timeline progression rate summary statistics, and 5 x 10^-8^ for AD case-control status^13^.

### Miscellaneous

Data processing and statistical analyses were primarily conducted using R (version 4.2.0 unless otherwise specified)^154^ and Python (version 3.12.10). Many R packages, including some from Bioconductor^155,156^, were used including “data.table” (v.1.18.4) for efficient reading and writing of large datasets; “tidyverse” (v.2.0.0)^157^ for data wrangling; “ggplot2” (v.4.0.3)^158^ for visualization; “patchwork” (v.1.3.2) for figure assembly; and “openxlsx” (v.4.2.8.1) for creating supplementary tables. Standard correlation tests (Spearman, Pearson, and Kendall) were conducted using the “cor.test()” R function. Linear models were fit with the “lm()” R function. Benjamini-Hochberg (FDR) corrections for multi-testing were performed unless otherwise specified using the “p.adjust()” R function. Genomic inflation (λ) was calculated with the “qchisq()” R function with the following R code: “median(qchisq(df$P, df = 1, lower.tail = FALSE), na.rm = TRUE) / qchisq(0.5, df = 1)”. Workflow management was conducted using Snakemake (v.9.3.0)^159^ with batch job submission to the Load Sharing Facility (LSF) queue using the snakemake executor lsf plugin (github.com/BEFH/snakemake-executor-plugin-lsf). Binary columns in supplementary tables were encoded such that “TRUE” is represented by “1” and “FALSE” is represented by “0” unless otherwise specified. Any p-values smaller than the minimum positive normalized double-precision floating point value (2^−1022^ ≈ 2.225074 × 10^−308^) are replaced with “< 2.23e-308” in supplementary tables.

## Data and Code Availability

All data used in the preparation of this article are publicly available under the regulation of each of the respective cohorts listed below.

The following data was obtained from the Alzheimer’s Disease Neuroimaging Initiative (ADNI) database (adni.loni.usc.edu). 1) Serum metabolites measured by FIA was obtained from the “ADMC Duke Biocrates MxP Quant 500 FIA Longitudinal [ADNI1,GO,2]” file in LONI-IDA on September 5, 2024 (synapse: syn52050341). 2) Serum metabolites measured by UPLC was obtained from the “ADMC Duke Biocrates MxP Quant 500 UPLC Longitudinal [ADNI1,GO,2]” file in LONI-IDA on September 5, 2024 (synapse: syn52050343). 3) Serum lipids and lipoproteins measured by NMR was obtained from the “ADMC Nightingale Platform NMR Analysis of Lipoproteins and Metabolites Longitudinal[ADNI1,GO,2]” file in LONI-IDA on September 5, 2024 (synapse: syn25985662). 4) Plasma lipids measured by LCMS was obtained from the “ADMC Lipidomics Meikle Lab Longitudinal Data Matrix [ADNI1,GO,2]” file in LONI-IDA on September 5, 2024 (synapse: syn52045871). 5) Serum metabolites measured using the METABOLON platform was obtained from the “ADMC Duke Metabolon DiscoveryHD4 Untargeted Serum (UPLC-MS/MS) [ADNI1,GO,2]” file in LONI-IDA on October 22, 2025 (synapse: syn70784221). 6) FIA and UPLC quality control data sheets was obtained from the “ADMC Duke Biocrates MxP Quant 500 FIA UPLC Longitudinal [ADNI1,GO,2] QC (Excel)” file in LONI-IDA on November 3, 2025 (synapse: syn71703725). 7) ADNI data dictionary was obtained from the “Data Dictionary [ADNI1,GO,2,3,4]” file in LONI-IDA on January 20, 2026. 8) APOE genotype information was obtained from the “ApoE Genotyping - Results [ADNI1,GO,2,3,4]” file in LONI-IDA on January 26, 2026. 9) ADNI1 genetic data was obtained from the “ADNI 1 SNP genotype data – PLINK” file in LONI-IDA on March 3, 2012. 10) ADNI2/GO genetic data was obtained from the “ADNI GO/2 SNP genotype data - Complete PLINK for sets 1 – 9” and “ADNI GO/2 SNP genotype data - Complete PLINK for sets 10 – 15” files in LONI-IDA on June 23, 2012 and July 17, 2014 respectively. 11) Plasma NfL biomarker measurements were obtained from the “Blennow Lab ADNI1-2 Plasma neurofilament light (NFL) longitudinal [ADNI1,GO,2]” file in LONI-IDA on August 26, 2025. 12) Plasma p-tau217, Aβ42, Aβ40, and Aβ42/Aβ40 ratio biomarker measurements were obtained from the “UPENN - Plasma Biomarkers (AB42, AB40, ptau217, NfL, GFAP) measured by Fujirebio & Quanterix [ADNI1,GO,2,3,4]” file in LONI-IDA on August 26, 2025. 13) The ADNIMERGE package was obtained from the “ADNIMERGE - Key ADNI tables merged into one Packages for R [ADNI1,GO,2,3,4]” in LONI-IDA on August 29, 2024.

The following summary statistics (identified by GWAS catalog accession ID) were obtained from the GWAS catalog (ebi.ac.uk/gwas) on May 12, 2026: “GCST002245”, “GCST005920”, “GCST005921”, “GCST005923”, “GCST011943”, “GCST007320”, “GCST90276158”, “GCST90475484”, “GCST90476375”, “GCST90475249”, “GCST90475496”, “GCST90475542”, “GCST90475543”, “GCST90475550”, “GCST90012877”, “GCST90012878”, “GCST90027158”, “GCST90435837”, “GCST90444478”, “GCST90421075”, “GCST90129599”, “GCST90129600”, “GCST90134626”, “GCST90134627”, “GCST90134628”, “GCST90134629”, “GCST90134630”, “GCST90134631”, “GCST90134632”, “GCST90134633”, “GCST90060018”, “GCST90470024”, “GCST90042678”, “GCST90042691”, “GCST90078284”, “GCST90079783”, “GCST90079819”, “GCST90078269”, and “GCST90078253”. GWAS catalog summary statistics are publicly available for download.

The following data was obtained from the Alzheimer’s Disease Sequencing Project (ADSP) and NIAGADS Data Sharing Service (https://dss.niagads.org). 1) Per-chromosome VCF genotype files were obtained from “s3://wanglab-dss-tier0/distribution/adsp/genotype/ALL/fsa000026/compact_filtered_multiallelic/gcad.preview.comp act_filtered.r4.wgs.36361.GATK.2022.08.15.multiallelic.genotypes.chr{1-22}.ALL.vcf.bgz” on September 26, 2024. 2) Case-control summary statistics were obtained frim the “GCST90027158_buildGRCh38.tsv.gz” file on July 25, 2024. 3)

All results and summary statistics presented in this article are either contained within associated supplementary tables or will be made publicly available in the GWAS catalog upon publication. All code associated with this manuscript will be made publicly available upon publication.

## Acknowledgments

### ADNI

Data collection and sharing for the Alzheimer’s Disease Neuroimaging Initiative (ADNI) is funded by the National Institute on Aging (National Institutes of Health Grant U19 AG024904). The grantee organization is the Northern California Institute for Research and Education. In the past, ADNI has also received funding from the National Institute of Biomedical Imaging and Bioengineering, the Canadian Institutes of Health Research, and private sector contributions through the Foundation for the National Institutes of Health (FNIH) including generous contributions from the following: AbbVie, Alzheimer’s Association; Alzheimer’s Drug Discovery Foundation; Araclon Biotech; BioClinica, Inc.; Biogen; Bristol-Myers Squibb Company; CereSpir, Inc.; Cogstate; Eisai Inc.; Elan Pharmaceuticals, Inc.; Eli Lilly and Company; EuroImmun; F. Hoffmann-La Roche Ltd and its affiliated company Genentech, Inc.; Fujirebio; GE Healthcare; IXICO Ltd.; Janssen Alzheimer Immunotherapy Research & Development, LLC.; Johnson & Johnson Pharmaceutical Research &Development LLC.; Lumosity; Lundbeck; Merck & Co., Inc.; Meso Scale Diagnostics, LLC.; NeuroRx Research; Neurotrack Technologies; Novartis Pharmaceuticals Corporation; Pfizer Inc.; Piramal Imaging; Servier; Takeda Pharmaceutical Company; and Transition Therapeutics.

### ADMC

Data collection and sharing for this project was funded by the Alzheimer’s Disease Metabolomics Consortium (National Institute on Aging R01AG046171, RF1AG051550 and 3U01AG024904-09S4).

### NIAGADS

Data for this study were prepared, archived, and distributed by the National Institute on Aging Alzheimer’s Disease Data Storage Site (NIAGADS) at the University of Pennsylvania (U24-AG041689), funded by the National Institute on Aging.

### Minerva

This work was supported in part through the Minerva computational and data resources and staff expertise provided by Scientific Computing and Data at the Icahn School of Medicine at Mount Sinai and supported by the Clinical and Translational Science Awards (CTSA) grant UL1TR004419 from the National Center for Advancing Translational Sciences.

### ADSP

Data for this study were prepared, archived, and distributed by the National Institute on Aging Alzheimer’s Disease Data Storage Site (NIAGADS) at the University of Pennsylvania (U24-AG041689), funded by the National Institute on Aging. Below we provide acknowledgments for the specific datasets in the ADSP release 4 whole genome sequencing data. We accessed these acknowledgments at https://dss.niagads.org/datasets/ng00067/ on May 1, 2026.

### Alzheimer’s Disease Sequencing Project (sa000001) data

The Alzheimer’s Disease Sequencing Project (ADSP) is comprised of two Alzheimer’s Disease (AD) genetics consortia and three National Human Genome Research Institute (NHGRI) funded Large Scale Sequencing and Analysis Centers (LSAC). The two AD genetics consortia are the Alzheimer’s Disease Genetics Consortium (ADGC) funded by NIA (U01 AG032984), and the Cohorts for Heart and Aging Research in Genomic Epidemiology (CHARGE) funded by NIA (R01 AG033193), the National Heart, Lung, and Blood Institute (NHLBI), other National Institute of Health (NIH) institutes and other foreign governmental and non-governmental organizations. The Discovery Phase analysis of sequence data is supported through UF1AG047133 (to Drs. Schellenberg, Farrer, Pericak-Vance, Mayeux, and Haines); U01AG049505 to Dr. Seshadri; U01AG049506 to Dr. Boerwinkle; U01AG049507 to Dr. Wijsman; and U01AG049508 to Dr. Goate and the Discovery Extension Phase analysis is supported through U01AG052411 to Dr. Goate, U01AG052410 to Dr. Pericak-Vance and U01 AG052409 to Drs. Seshadri and Fornage.

Sequencing for the Follow Up Study (FUS) is supported through U01AG057659 (to Drs. PericakVance, Mayeux, and Vardarajan) and U01AG062943 (to Drs. Pericak-Vance and Mayeux). Data generation and harmonization in the Follow-up Phase is supported by U54AG052427 (to Drs. Schellenberg and Wang). The FUS Phase analysis of sequence data is supported through U01AG058589 (to Drs. Destefano, Boerwinkle, De Jager, Fornage, Seshadri, and Wijsman), U01AG058654 (to Drs. Haines, Bush, Farrer, Martin, and Pericak-Vance), U01AG058635 (to Dr. Goate), RF1AG058066 (to Drs. Haines, Pericak-Vance, and Scott), RF1AG057519 (to Drs. Farrer and Jun), R01AG048927 (to Dr. Farrer), and RF1AG054074 (to Drs. Pericak-Vance and Beecham).

The ADGC cohorts include: Adult Changes in Thought (ACT) (U01 AG006781, U19 AG066567), the Alzheimer’s Disease Research Centers (ADRC) (P30 AG062429, P30 AG066468, P30 AG062421, P30 AG066509, P30 AG066514, P30 AG066530, P30 AG066507, P30 AG066444, P30 AG066518, P30 AG066512, P30 AG066462, P30 AG072979, P30 AG072972, P30 AG072976, P30 AG072975, P30 AG072978, P30 AG072977, P30 AG066519, P30 AG062677, P30 AG079280, P30 AG062422, P30 AG066511, P30 AG072946, P30 AG062715, P30 AG072973, P30 AG066506, P30 AG066508, P30 AG066515, P30 AG072947, P30 AG072931, P30 AG066546, P20 AG068024, P20 AG068053, P20 AG068077, P20 AG068082, P30 AG072958, P30 AG072959), the Chicago Health and Aging Project (CHAP) (R01 AG11101, RC4 AG039085, K23 AG030944), Indiana Memory and Aging Study (IMAS) (R01 AG019771), Indianapolis Ibadan (R01 AG009956, P30 AG010133), the Memory and Aging Project (MAP) (R01 AG17917), Mayo Clinic (MAYO) (R01 AG032990, U01 AG046139, R01 NS080820, RF1 AG051504, P50 AG016574), Mayo Parkinson’s Disease controls (NS039764, NS071674, 5RC2HG005605), University of Miami (R01 AG027944, R01 AG028786, R01 AG019085, IIRG09133827, A2011048), the Multi-Institutional Research in Alzheimer’s Genetic Epidemiology Study (MIRAGE) (R01 AG09029, R01 AG025259), the National Centralized Repository for Alzheimer’s Disease and Related Dementias (NCRAD) (U24 AG021886), the National Institute on Aging Late Onset Alzheimer’s Disease Family Study (NIA- LOAD) (U24 AG056270), the Religious Orders Study (ROS) (P30 AG10161, R01 AG15819), the Texas Alzheimer’s Research and Care Consortium (TARCC) (funded by the Darrell K Royal Texas Alzheimer’s Initiative), Vanderbilt University/Case Western Reserve University (VAN/CWRU) (R01 AG019757, R01 AG021547, R01 AG027944, R01 AG028786, P01 NS026630, and Alzheimer’s Association), the Washington Heights-Inwood Columbia Aging Project (WHICAP) (RF1 AG054023), the University of Washington Families (VA Research Merit Grant, NIA: P50AG005136, R01AG041797, NINDS: R01NS069719), the Columbia University Hispanic Estudio Familiar de Influencia Genetica de Alzheimer (EFIGA) (RF1 AG015473), the University of Toronto (UT) (funded by Wellcome Trust, Medical Research Council, Canadian Institutes of Health Research), and Genetic Differences (GD) (R01 AG007584). The CHARGE cohorts are supported in part by National Heart, Lung, and Blood Institute (NHLBI) infrastructure grant HL105756 (Psaty), RC2HL102419 (Boerwinkle) and the neurology working group is supported by the National Institute on Aging (NIA) R01 grant AG033193.

The CHARGE cohorts participating in the ADSP include the following: Austrian Stroke Prevention Study (ASPS), ASPS-Family study, and the Prospective Dementia Registry-Austria (ASPS/PRODEM-Aus), the Atherosclerosis Risk in Communities (ARIC) Study, the Cardiovascular Health Study (CHS), the Erasmus Rucphen Family Study (ERF), the Framingham Heart Study (FHS), and the Rotterdam Study (RS). ASPS is funded by the Austrian Science Fond (FWF) grant number P20545-P05 and P13180 and the Medical University of Graz. The ASPS-Fam is funded by the Austrian Science Fund (FWF) project I904), the EU Joint Programme – Neurodegenerative Disease Research (JPND) in frame of the BRIDGET project (Austria, Ministry of Science) and the Medical University of Graz and the Steiermärkische Krankenanstalten Gesellschaft. PRODEM-Austria is supported by the Austrian Research Promotion agency (FFG) (Project No. 827462) and by the Austrian National Bank (Anniversary Fund, project 15435. ARIC research is carried out as a collaborative study supported by NHLBI contracts (HHSN268201100005C, HHSN268201100006C, HHSN268201100007C, HHSN268201100008C, HHSN268201100009C, HHSN268201100010C, HHSN268201100011C, and HHSN268201100012C). Neurocognitive data in ARIC is collected by U01 2U01HL096812, 2U01HL096814, 2U01HL096899, 2U01HL096902, 2U01HL096917 from the NIH (NHLBI, NINDS, NIA and NIDCD), and with previous brain MRI examinations funded by R01-HL70825 from the NHLBI. CHS research was supported by contracts HHSN268201200036C, HHSN268200800007C, N01HC55222, N01HC85079, N01HC85080, N01HC85081, N01HC85082, N01HC85083, N01HC85086, and grants U01HL080295 and U01HL130114 from the NHLBI with additional contribution from the National Institute of Neurological Disorders and Stroke (NINDS). Additional support was provided by R01AG023629, R01AG15928, and R01AG20098 from the NIA. FHS research is supported by NHLBI contracts N01-HC-25195 and HHSN268201500001I. This study was also supported by additional grants from the NIA (R01s AG054076, AG049607 and AG033040 and NINDS (R01 NS017950). The ERF study as a part of EUROSPAN (European Special Populations Research Network) was supported by European Commission FP6 STRP grant number 018947 (LSHG-CT-2006-01947) and also received funding from the European Community’s Seventh Framework Programme (FP7/2007-2013)/grant agreement HEALTH-F4- 2007-201413 by the European Commission under the programme “Quality of Life and Management of the Living Resources” of 5th Framework Programme (no. QLG2-CT-2002- 01254). High-throughput analysis of the ERF data was supported by a joint grant from the Netherlands Organization for Scientific Research and the Russian Foundation for Basic Research (NWO-RFBR 047.017.043). The Rotterdam Study is funded by Erasmus Medical Center and Erasmus University, Rotterdam, the Netherlands Organization for Health Research and Development (ZonMw), the Research Institute for Diseases in the Elderly (RIDE), the Ministry of Education, Culture and Science, the Ministry for Health, Welfare and Sports, the European Commission (DG XII), and the municipality of Rotterdam. Genetic data sets are also supported by the Netherlands Organization of Scientific Research NWO Investments (175.010.2005.011, 911-03-012), the Genetic Laboratory of the Department of Internal Medicine, Erasmus MC, the Research Institute for Diseases in the Elderly (014-93-015; RIDE2), and the Netherlands Genomics Initiative (NGI)/Netherlands Organization for Scientific Research (NWO) Netherlands Consortium for Healthy Aging (NCHA), project 050-060-810. All studies are grateful to their participants, faculty and staff. The content of these manuscripts is solely the responsibility of the authors and does not necessarily represent the official views of the National Institutes of Health or the U.S. Department of Health and Human Services.

The FUS cohorts include: the Alzheimer’s Disease Research Centers (ADRC) (P30 AG062429, P30 AG066468, P30 AG062421, P30 AG066509, P30 AG066514, P30 AG066530, P30 AG066507, P30 AG066444, P30 AG066518, P30 AG066512, P30 AG066462, P30 AG072979, P30 AG072972, P30 AG072976, P30 AG072975, P30 AG072978, P30 AG072977, P30 AG066519, P30 AG062677, P30 AG079280, P30 AG062422, P30 AG066511, P30 AG072946, P30 AG062715, P30 AG072973, P30 AG066506, P30 AG066508, P30 AG066515, P30 AG072947, P30 AG072931, P30 AG066546, P20 AG068024, P20 AG068053, P20 AG068077, P20 AG068082, P30 AG072958, P30 AG072959), Alzheimer’s Disease Neuroimaging Initiative (ADNI) (U19AG024904), Amish Protective Variant Study (RF1AG058066), Cache County Study (R01AG11380, R01AG031272, R01AG21136, RF1AG054052), Case Western Reserve University Brain Bank (CWRUBB) (P50AG008012), Case Western Reserve University Rapid Decline (CWRURD) (RF1AG058267, NU38CK000480), CubanAmerican Alzheimer’s Disease Initiative (CuAADI) (3U01AG052410), Estudio Familiar de Influencia Genetica en Alzheimer (EFIGA) (5R37AG015473, RF1AG015473, R56AG051876), Genetic and Environmental Risk Factors for Alzheimer Disease Among African Americans Study (GenerAAtions) (2R01AG09029, R01AG025259, 2R01AG048927), Gwangju Alzheimer and Related Dementias Study (GARD) (U01AG062602), Hillblom Aging Network (2014-A-004-NET, R01AG032289, R01AG048234), Hussman Institute for Human Genomics Brain Bank (HIHGBB) (R01AG027944, Alzheimer’s Association “Identification of Rare Variants in Alzheimer Disease”), Ibadan Study of Aging (IBADAN) (5R01AG009956), Longevity Genes Project (LGP) and LonGenity (R01AG042188, R01AG044829, R01AG046949, R01AG057909, R01AG061155, P30AG038072), Mexican Health and Aging Study (MHAS) (R01AG018016), Multi-Institutional Research in Alzheimer’s Genetic Epidemiology (MIRAGE) (2R01AG09029, R01AG025259, 2R01AG048927), Northern Manhattan Study (NOMAS) (R01NS29993), Peru Alzheimer’s Disease Initiative (PeADI) (RF1AG054074), Puerto Rican 1066 (PR1066) (Wellcome Trust (GR066133/GR080002), European Research Council (340755)), Puerto Rican Alzheimer Disease Initiative (PRADI) (RF1AG054074), Reasons for Geographic and Racial Differences in Stroke (REGARDS) (U01NS041588), Research in African American Alzheimer Disease Initiative (REAAADI) (U01AG052410), the Religious Orders Study (ROS) (P30 AG10161, P30 AG72975, R01 AG15819, R01 AG42210), the RUSH Memory and Aging Project (MAP) (R01 AG017917, R01 AG42210Stanford Extreme Phenotypes in AD (R01AG060747), University of Miami Brain Endowment Bank (MBB), University of Miami/Case Western/North Carolina A&T African American (UM/CASE/NCAT) (U01AG052410, R01AG028786), Wisconsin Registry for Alzheimer’s Prevention (WRAP) (R01AG027161 and R01AG054047), Mexico-Southern California Autosomal Dominant Alzheimer’s Disease Consortium (R01AG069013), Center for Cognitive Neuroscience and Aging (R01AG047649), and the A4 Study (R01AG063689, U19AG010483 and U24AG057437).

The four LSACs are: the Human Genome Sequencing Center at the Baylor College of Medicine (U54 HG003273), the Broad Institute Genome Center (U54HG003067), The American Genome Center at the Uniformed Services University of the Health Sciences (U01AG057659), and the Washington University Genome Institute (U54HG003079). Genotyping and sequencing for the ADSP FUS is also conducted at John P. Hussman Institute for Human Genomics (HIHG) Center for Genome Technology (CGT).

Biological samples and associated phenotypic data used in primary data analyses were stored at Study Investigators institutions, and at the National Centralized Repository for Alzheimer’s Disease and Related Dementias (NCRAD, U24AG021886) at Indiana University funded by NIA. Associated Phenotypic Data used in primary and secondary data analyses were provided by Study Investigators, the NIA funded Alzheimer’s Disease Centers (ADCs), and the National Alzheimer’s Coordinating Center (NACC, U24AG072122) and the National Institute on Aging Genetics of Alzheimer’s Disease Data Storage Site (NIAGADS, U24AG041689) at the University of Pennsylvania, funded by NIA. Harmonized phenotypes were provided by the ADSP Phenotype Harmonization Consortium (ADSP-PHC), funded by NIA (U24 AG074855, U01 AG068057 and R01 AG059716) and Ultrascale Machine Learning to Empower Discovery in Alzheimer’s Disease Biobanks (AI4AD, U01 AG068057). This research was supported in part by the Intramural Research Program of the National Institutes of health, National Library of Medicine. Contributors to the Genetic Analysis Data included Study Investigators on projects that were individually funded by NIA, and other NIH institutes, and by private U.S. organizations, or foreign governmental or nongovernmental organizations.

The ADSP Phenotype Harmonization Consortium (ADSP-PHC) is funded by NIA (U24 AG074855, U01 AG068057 and R01 AG059716). The harmonized cohorts within the ADSP-PHC include: the Anti-Amyloid Treatment in Asymptomatic Alzheimer’s study (A4 Study), a secondary prevention trial in preclinical Alzheimer’s disease, aiming to slow cognitive decline associated with brain amyloid accumulation in clinically normal older individuals. The A4 Study is funded by a public-private-philanthropic partnership, including funding from the National Institutes of Health-National Institute on Aging, Eli Lilly and Company, Alzheimer’s Association, Accelerating Medicines Partnership, GHR Foundation, an anonymous foundation and additional private donors, with in-kind support from Avid and Cogstate. The companion observational Longitudinal Evaluation of Amyloid Risk and Neurodegeneration (LEARN) Study is funded by the Alzheimer’s Association and GHR Foundation. The A4 and A4-LEARN Studies are led by Dr. Reisa Sperling at Brigham and Women’s Hospital, Harvard Medical School and Dr. Paul Aisen at the Alzheimer’s Therapeutic Research Institute (ATRI), University of Southern California. The A4 and LEARN Studies are coordinated by ATRI at the University of Southern California, and the data are made available through the Laboratory for Neuro Imaging at the University of Southern California. The participants screening for the A4 Study provided permission to share their de-identified data in order to advance the quest to find a successful treatment for Alzheimer’s disease. We would like to acknowledge the dedication of all the participants, the site personnel, and all of the partnership team members who continue to make the A4 and LEARN Studies possible. The complete A4 Study Team list is available on: https://a4study.org/a4-study-team.; the Adult Changes in Thought study (ACT), U01 AG006781, U19 AG066567; Alzheimer’s Disease Neuroimaging Initiative (ADNI): Data collection and sharing for this project was funded by the Alzheimer’s Disease Neuroimaging Initiative (ADNI) (National Institutes of Health Grant U01 AG024904) and DOD ADNI (Department of Defense award number W81XWH-12-2-0012). ADNI is funded by the National Institute on Aging, the National Institute of Biomedical Imaging and Bioengineering, and through generous contributions from the following: AbbVie, Alzheimer’s Association; Alzheimer’s Drug Discovery Foundation; Araclon Biotech; BioClinica, Inc.; Biogen; Bristol-Myers Squibb Company; CereSpir, Inc.; Cogstate; Eisai Inc.; Elan Pharmaceuticals, Inc.; Eli Lilly and Company; EuroImmun; F. Hoffmann-La Roche Ltd and its affiliated company Genentech, Inc.; Fujirebio; GE Healthcare; IXICO Ltd.; Janssen Alzheimer Immunotherapy Research & Development, LLC.; Johnson & Johnson Pharmaceutical Research & Development LLC.; Lumosity; Lundbeck; Merck & Co., Inc.;Meso Scale Diagnostics, LLC.; NeuroRx Research; Neurotrack Technologies; Novartis Pharmaceuticals Corporation; Pfizer Inc.; Piramal Imaging; Servier; Takeda Pharmaceutical Company; and Transition Therapeutics. The Canadian Institutes of Health Research is providing funds to support ADNI clinical sites in Canada. Private sector contributions are facilitated by the Foundation for the National Institutes of Health (https://www.fnih.org). The grantee organization is the Northern California Institute for Research and Education, and the study is coordinated by the Alzheimer’s Therapeutic Research Institute at the University of Southern California. ADNI data are disseminated by the Laboratory for Neuro Imaging at the University of Southern California; Brain tissue and additional biological samples were collected from individuals who were assessed and followed in the Case Western ADC which was funded by NIA P50AG008012; Estudio Familiar de Influencia Genetica en Alzheimer (EFIGA): 5R37AG015473, RF1AG015473, R56AG051876; the Health & Aging Brain Study – Health Disparities (HABS-HD), supported by the National Institute on Aging of the National Institutes of Health under Award Numbers R01AG054073, R01AG058533, R01AG070862, P41EB015922, and U19AG078109; brain tissues were obtained from the Brain Bank at the John P. Hussman Institute for Human Genomics at the University of Miami Miller School of Medicine, with funding provided by Louis D. Scientific Prize from the Institut de France; the Indiana Alzheimer’s Disease Research Center (IADRC) supported by National Institutes of Health (NIH) grant P30AG072976; the Korean Brain Aging Study for the Early Diagnosis and Prediction of Alzheimer’s disease (KBASE), which was supported by a grant from Ministry of Science, ICT and Future Planning (Grant No: NRF-2014M3C7A1046042); Memory & Aging Project at Knight Alzheimer’s Disease Research Center (MAP at Knight ADRC): The Memory and Aging Project at the Knight-ADRC (Knight-ADRC). This work was supported by the National Institutes of Health (NIH) grants R01AG064614, R01AG044546, RF1AG053303, RF1AG058501, U01AG058922 and R01AG064877 to Carlos Cruchaga. The recruitment and clinical characterization of research participants at Washington University was supported by NIH grants P30AG066444, P01AG03991, and P01AG026276. Data collection and sharing for this project was supported by NIH grants RF1AG054080, P30AG066462, R01AG064614 and U01AG052410. We thank the contributors who collected samples used in this study, as well as patients and their families, whose help and participation made this work possible. This work was supported by access to equipment made possible by the Hope Center for Neurological Disorders, the Neurogenomics and Informatics Center (NGI: https://neurogenomics.wustl.edu/) and the Departments of Neurology and Psychiatry at Washington University School of Medicine; the University of Miami Brain Endowment Bank (Miami Brain Bank); National Alzheimer’s Coordinating Center (NACC): The NACC database is funded by NIA/NIH Grant U24 AG072122. SCAN is a multi-institutional project that was funded as a U24 grant (AG067418) by the National Institute on Aging in May 2020. Data collected by SCAN and shared by NACC are contributed by the NIA-funded ADRCs as follows: P30 AG062429 (PI James Brewer, MD, PhD), P30 AG066468 (PI Oscar Lopez, MD), P30 AG062421 (PI Bradley Hyman, MD, PhD), P30 AG066509 (PI Thomas Grabowski, MD), P30 AG066514 (PI Mary Sano, PhD), P30 AG066530 (PI Helena Chui, MD), P30 AG066507 (PI Marilyn Albert, PhD), P30 AG066444 (PI John Morris, MD), P30 AG066518 (PI Jeffrey Kaye, MD), P30 AG066512 (PI Thomas Wisniewski, MD), P30 AG066462 (PI Scott Small, MD), P30 AG072979 (PI David Wolk, MD), P30 AG072972 (PI Charles DeCarli, MD), P30 AG072976 (PI Andrew Saykin, PsyD), P30 AG072975 (PI David Bennett, MD), P30 AG072978 (PI Neil Kowall, MD), P30 AG072977 (PI Robert Vassar, PhD), P30 AG066519 (PI Frank LaFerla, PhD), P30 AG062677 (PI Ronald Petersen, MD, PhD), P30 AG079280 (PI Eric Reiman, MD), P30 AG062422 (PI Gil Rabinovici, MD), P30 AG066511 (PI Allan Levey, MD, PhD), P30 AG072946 (PI Linda Van Eldik, PhD), P30 AG062715 (PI Sanjay Asthana, MD, FRCP), P30 AG072973 (PI Russell Swerdlow, MD), P30 AG066506 (PI Todd Golde, MD, PhD), P30 AG066508 (PI Stephen Strittmatter, MD, PhD), P30 AG066515 (PI Victor Henderson, MD, MS), P30 AG072947 (PI Suzanne Craft, PhD), P30 AG072931 (PI Henry Paulson, MD, PhD), P30 AG066546 (PI Sudha Seshadri, MD), P20 AG068024 (PI Erik Roberson, MD, PhD), P20 AG068053 (PI Justin Miller, PhD), P20 AG068077 (PI Gary Rosenberg, MD), P20 AG068082 (PI Angela Jefferson, PhD), P30 AG072958 (PI Heather Whitson, MD), P30 AG072959 (PI James Leverenz, MD); National Institute on Aging Alzheimer’s Disease Family Based Study (NIA-AD FBS): U24 AG056270; Religious Orders Study (ROS): P30AG10161,R01AG15819, R01AG42210; Memory and Aging Project (MAP - Rush): R01AG017917, R01AG42210; Minority Aging Research Study (MARS): R01AG22018, R01AG42210; the Texas Alzheimer’s Research and Care Consortium (TARCC), funded by the Darrell K Royal Texas Alzheimer’s Initiative, directed by the Texas Council on Alzheimer’s Disease and Related Disorders; Washington Heights/Inwood Columbia Aging Project (WHICAP): RF1 AG054023;and Wisconsin Registry for Alzheimer’s Prevention (WRAP): R01AG027161 and R01AG054047. Additional acknowledgments include the National Institute on Aging Genetics of Alzheimer’s Disease Data Storage Site (NIAGADS, U24AG041689) at the University of Pennsylvania, funded by NIA. Last Updated 03.11.2026

### For investigators using Alzheimer’s Disease Neuroimaging Initiative (sa000002) data

Data collection and sharing for this project was funded by the Alzheimer’s Disease Neuroimaging Initiative (ADNI) (National Institutes of Health Grant U01 AG024904) and DOD ADNI (Department of Defense award number W81XWH-12-2-0012). ADNI is funded by the National Institute on Aging, the National Institute of Biomedical Imaging and Bioengineering, and through generous contributions from the following: AbbVie, Alzheimer’s Association; Alzheimer’s Drug Discovery Foundation; Araclon Biotech; BioClinica, Inc.; Biogen; Bristol-Myers Squibb Company; CereSpir, Inc.; Cogstate; Eisai Inc.; Elan Pharmaceuticals, Inc.; Eli Lilly and Company; EuroImmun; F. Hoffmann-La Roche Ltd and its affiliated company Genentech, Inc.; Fujirebio; GE Healthcare; IXICO Ltd.; Janssen Alzheimer Immunotherapy Research & Development, LLC.; Johnson & Johnson Pharmaceutical Research & Development LLC.; Lumosity; Lundbeck; Merck & Co., Inc.; Meso Scale Diagnostics, LLC.; NeuroRx Research; Neurotrack Technologies; Novartis Pharmaceuticals Corporation; Pfizer Inc.; Piramal Imaging; Servier; Takeda Pharmaceutical Company; and Transition Therapeutics. The Canadian Institutes of Health Research is providing funds to support ADNI clinical sites in Canada. Private sector contributions are facilitated by the Foundation for the National Institutes of Health (https://www.fnih.org). The grantee organization is the Northern California Institute for Research and Education, and the study is coordinated by the Alzheimer’s Therapeutic Research Institute at the University of Southern California. ADNI data are disseminated by the Laboratory for Neuro Imaging at the University of Southern California. Additional information to include in an acknowledgment statement can be found on the LONI site: https://adni.loni.usc.edu/wp-content/uploads/how_to_apply/ADNI_Data_Use_Agreement.pdf.

### For investigators using Alzheimer’s Disease Genetics Consortium (sa000003) data

The Alzheimer’s Disease Genetics Consortium (ADGC) supported sample preparation, sequencing and data processing through NIA grant U01AG032984. Sequencing data generation and harmonization is supported by the Genome Center for Alzheimer’s Disease, U54AG052427, and data sharing is supported by NIAGADS, U24AG041689. Samples from the National Centralized Repository for Alzheimer’s Disease and Related Dementias (NCRAD), which receives government support under a cooperative agreement grant (U24 AG021886) awarded by the National Institute on Aging (NIA), were used in this study. We thank contributors who collected samples used in this study, as well as patients and their families, whose help and participation made this work possible.

### GWAS Datasets ADC1-15

The NACC database is funded by NIA/NIH Grant U24 AG072122. NACC data are contributed by the NIA-funded ADRCs: P30 AG062429 (PI James Brewer, MD, PhD), P30 AG066468 (PI Oscar Lopez, MD), P30 AG062421 (PI Bradley Hyman, MD, PhD), P30 AG066509 (PI Thomas Grabowski, MD), P30 AG066514 (PI Mary Sano, PhD), P30 AG066530 (PI Helena Chui, MD), P30 AG066507 (PI Marilyn Albert, PhD), P30 AG066444 (PI John Morris, MD), P30 AG066518 (PI Jeffrey Kaye, MD), P30 AG066512 (PI Thomas Wisniewski, MD), P30 AG066462 (PI Scott Small, MD), P30 AG072979 (PI David Wolk, MD), P30 AG072972 (PI Charles DeCarli, MD), P30 AG072976 (PI Andrew Saykin, PsyD), P30 AG072975 (PI David Bennett, MD), P30 AG072978 (PI Neil Kowall, MD), P30 AG072977 (PI Robert Vassar, PhD), P30 AG066519 (PI Frank LaFerla, PhD), P30 AG062677 (PI Ronald Petersen, MD, PhD), P30 AG079280 (PI Eric Reiman, MD), P30 AG062422 (PI Gil Rabinovici, MD), P30 AG066511 (PI Allan Levey, MD, PhD), P30 AG072946 (PI Linda Van Eldik, PhD), P30 AG062715 (PI Sanjay Asthana, MD, FRCP), P30 AG072973 (PI Russell Swerdlow, MD), P30 AG066506 (PI Todd Golde, MD, PhD), P30 AG066508 (PI Stephen Strittmatter, MD, PhD), P30 AG066515 (PI Victor Henderson, MD, MS), P30 AG072947 (PI Suzanne Craft, PhD), P30 AG072931 (PI Henry Paulson, MD, PhD), P30 AG066546 (PI Sudha Seshadri, MD), P20 AG068024 (PI Erik Roberson, MD, PhD), P20 AG068053 (PI Justin Miller, PhD), P20 AG068077 (PI Gary Rosenberg, MD), P20 AG068082 (PI Angela Jefferson, PhD), P30 AG072958 (PI Heather Whitson, MD), P30 AG072959 (PI James Leverenz, MD). NACC phenotypes were provided by the ADSP Phenotype Harmonization Consortium (ADSP-PHC), funded by NIA (U24 AG074855, U01 AG068057 and R01 AG059716).

### ADGC_AA_WES data

NIH grants supported enrollment and data collection for the individual studies including: GenerAAtions R01AG20688 (PI M. Daniele Fallin, PhD); Miami/Duke R01 AG027944, R01 AG028786 (PI Margaret A. Pericak-Vance, PhD); NC A&T P20 MD000546, R01 AG28786-01A1 (PI Goldie S. Byrd, PhD); Case Western (PI Jonathan L. Haines, PhD); MIRAGE R01 AG009029 (PI Lindsay A. Farrer, PhD); ROS P30AG10161, R01AG15819, R01AG30146, TGen (PI David A. Bennett, MD); MAP R01AG17917, R01AG15819, TGen (PI David A. Bennett, MD); MARS R01AG022018 (PI Lisa L. Barnes).[[CL1](https://dss.niagads.org/portal/darm/admin#_msocom_1)] [[KA2](https://dss.niagads.org/portal/darm/admin#_msocom_2)] The NACC database is funded by NIA/NIH Grant U24 AG072122. NACC data are contributed by the NIA-funded ADCs: P30 AG019610 (PI Eric Reiman, MD), P30 AG013846 (PI Neil Kowall, MD), P30 AG062428-01 (PI James Leverenz, MD) P50 AG008702 (PI Scott Small, MD), P50 AG025688 (PI Allan Levey, MD, PhD), P50 AG047266 (PI Todd Golde, MD, PhD), P30 AG010133 (PI Andrew Saykin, PsyD), P50 AG005146 (PI Marilyn Albert, PhD), P30 AG062421-01 (PI Bradley Hyman, MD, PhD), P30 AG062422-01 (PI Ronald Petersen, MD, PhD), P50 AG005138 (PI Mary Sano, PhD), P30 AG008051 (PI Thomas Wisniewski, MD), P30 AG013854 (PI Robert Vassar, PhD), P30 AG008017 (PI Jeffrey Kaye, MD), P30 AG010161 (PI David Bennett, MD), P50 AG047366 (PI Victor Henderson, MD, MS), P30 AG010129 (PI Charles DeCarli, MD), P50 AG016573 (PI Frank LaFerla, PhD), P30 AG062429-01(PI James Brewer, MD, PhD), P50 AG023501 (PI Bruce Miller, MD), P30 AG035982 (PI Russell Swerdlow, MD), P30 AG028383 (PI Linda Van Eldik, PhD), P30 AG053760 (PI Henry Paulson, MD, PhD), P30 AG010124 (PI John Trojanowski, MD, PhD), P50 AG005133 (PI Oscar Lopez, MD), P50 AG005142 (PI Helena Chui, MD), P30 AG012300 (PI Roger Rosenberg, MD), P30 AG049638 (PI Suzanne Craft, PhD), P50 AG005136 (PI Thomas Grabowski, MD), P30 AG062715-01 (PI Sanjay Asthana, MD, FRCP), P50 AG005681 (PI John Morris, MD), P50 AG047270 (PI Stephen Strittmatter, MD, PhD).

### ADGC-TARCC-WGS data

This study was made possible by the Texas Alzheimer’s Research and Care Consortium (TARCC) funded by the state of Texas through the Texas Council on Alzheimer’s Disease and Related Disorders and the Darrell K Royal Texas Alzheimer’s Initiative.

### The Familial Alzheimer Sequencing Project (sa000004) data

This work was supported by grants from the National Institutes of Health (R01AG044546, P01AG003991, RF1AG053303, R01AG058501, U01AG058922, RF1AG058501 and R01AG057777). The recruitment and clinical characterization of research participants at Washington University were supported by NIH P50 AG05681, P01 AG03991, and P01 AG026276. This work was supported by access to equipment made possible by the Hope Center for Neurological Disorders, and the Departments of Neurology and Psychiatry at Washington University School of Medicine. We thank the contributors who collected samples used in this study, as well as patients and their families, whose help and participation made this work possible. This work was supported by access to equipment made possible by the Hope Center for Neurological Disorders, and the Departments of Neurology and Psychiatry at Washington University School of Medicine.

### Brkanac- Family-based genome scan for AAO of LOAD (sa000005) data

This work was partially supported by grant funding from NIH R01 AG039700 and NIH P50 AG005136. Subjects and samples used here were originally collected with grant funding from NIH U24 AG026395, U24 AG021886, P50 AG008702, P01 AG007232, R37 AG015473, P30 AG028377, P50 AG05128, P50 AG16574, P30 AG010133, P50 AG005681, P01 AG003991, U01MH046281, U01 MH046290 and U01 MH046373. The funders had no role in study design, analysis or preparation of the manuscript. The authors declare no competing interests.

### HIHG Miami Families with AD (sa000006) data

This work was supported by the National Institutes of Health (R01 AG027944, R01 AG028786 to MAPV, R01 AG019085 to JLH, P20 MD000546); a joint grant from the Alzheimer’s Association (SG-14-312644) and the Fidelity Biosciences Research Initiative to MAPV; the BrightFocus Foundation (A2011048 to MAPV). NIA-LOAD Family-Based Study supported the collection of samples used in this study through NIH grants U24 AG026395 and R01 AG041797 and the MIRAGE cohort was supported through the NIH grants R01 AG025259 and R01 AG048927. We thank contributors, including the Alzheimer’s disease Centers who collected samples used in this study, as well as patients and their families, whose help and participation made this work possible. Study design: HNC, BWK, JLH, MAPV; Sample collection: MLC, JMV, RMC, LAF, JLH, MAPV; Whole exome sequencing and Sanger sequencing: SR, PLW; Sequencing data analysis: HNC, BWK, KLHN, SR, MAK, JRG, ERM, GWB, MAPV; Statistical analysis: BWK, KLHN, JMJ, MAPV; Preparation of manuscript: HNC, BWK. The authors jointly discussed the experimental results throughout the duration of the study. All authors read and approved the final manuscript.

### Washington Heights and Inwood Community Aging project (WHICAP) (sa000007) data

Data collection and sharing for this project was supported by the Washington Heights-Inwood Columbia Aging Project (WHICAP R01AG072474, PO1AG07232, R01AG037212, RF1AG054023) funded by the National Institute on Aging (NIA). This manuscript has been reviewed by WHICAP investigators for scientific content and consistency of data interpretation with previous WHICAP Study publications. We acknowledge the WHICAP study participants and the WHICAP research and support staff for their contributions to this study.

### Charles F. and Joanne Knight Alzheimer’s Disease Research Center (sa000008) data

This work was supported by grants from the National Institutes of Health (R01AG044546, P01AG003991, RF1AG053303, R01AG058501, U01AG058922, RF1AG058501 and R01AG057777). The recruitment and clinical characterization of research participants at Washington University were supported by NIH P50 AG05681, P01 AG03991, and P01 AG026276. This work was supported by access to equipment made possible by the Hope Center for Neurological Disorders, and the Departments of Neurology and Psychiatry at Washington University School of Medicine. We thank the contributors who collected samples used in this study, as well as patients and their families, whose help and participation made this work possible. This work was supported by access to equipment made possible by the Hope Center for Neurological Disorders, and the Departments of Neurology and Psychiatry at Washington University School of Medicine.

### ADSP-PHC harmonized phenotypes deposited within dataset, ng00067

The Memory and Aging Project at the Knight-ADRC (Knight-ADRC), supported by NIH grants R01AG064614, R01AG044546, RF1AG053303, RF1AG058501, U01AG058922 and R01AG064877 to Carlos Cruchaga. The recruitment and clinical characterization of research participants at Washington University was supported by NIH grants P30AG066444, P01AG03991, and P01AG026276. Data collection and sharing for this project was supported by NIH grants RF1AG054080, P30AG066462, R01AG064614 and U01AG052410. This work was supported by access to equipment made possible by the Hope Center for Neurological Disorders, the Neurogenomics and Informatics Center (NGI: https://neurogenomics.wustl.edu/) and the Departments of Neurology and Psychiatry at Washington University School of Medicine.

### ng00050 and ng00052 data

This work was supported by Pfizer and grants from the National Institutes of Health (R01-AG044546, P01-AG003991), and the Alzheimer’s Association (NIRG-11–200110). This research was conducted while Carlos Cruchaga was a recipient of a New Investigator Award in Alzheimer’s disease from the American Federation for Aging Research. Carlos Cruchaga is a recipient of a BrightFocus Foundation Alzheimer’s Disease Research Grant (A2013359S). The recruitment and clinical characterization of research participants at Washington University were supported by NIHP50 AG05681, P01 AG03991, and P01 AG026276. Some of the samples used in this study were genotyped by the ADGC and GERAD. ADGC is supported by grants from the NIH (#U01AG032984) and GERAD from the Wellcome Trust (GR082604MA) and the Medical Research Council (G0300429). Data collection and sharing for this project was funded by the Alzheimer’s Disease Neuroimaging Initiative (ADNI) (National Institutes of Health Grant U01 AG024904) and DOD ADNI (Department of Defense award number W81XWH-12-2-0012). ADNI is funded by the National Institute on Aging, the National Institute of Biomedical Imaging and Bioengineering, and through generous contributions from the following: Alzheimer’s Association; Alzheimer’s Drug Discovery Foundation; Araclon Biotech; BioClinica, Inc.; Biogen Idec; Bristol-Myers Squibb Company; Eisai; Elan Pharmaceuticals, Inc.; Eli Lilly and Company; EuroImmun; F. Hoffmann-La Roche Ltd. and its affiliated company Genentech, Inc.; Fujirebio; GE Healthcare; IXICO Ltd.; Janssen Alzheimer Immunotherapy Research & Development, LLC; Johnson & Johnson Pharmaceutical Research & Development LLC; Medpace; Merck; Meso Scale Diagnostics, LLC.; NeuroRx Research; Neurotrack Technologies; Novartis Pharmaceuticals Corporation; Pfizer Inc.; Piramal Imaging; Servier; Synarc Inc.; and Takeda Pharmaceutical Company. The Canadian Institutes of Rev December 5, 2013 Health Research is providing funds to support ADNI clinical sites in Canada. Private sector contributions are facilitated by the Foundation for the National Institutes of Health (https://www.fnih.org). The grantee organization is the Northern California Institute for Research and Education, and the study is coordinated by the Alzheimer’s Disease Cooperative Study at the University of California, San Diego. ADNI data are disseminated by the Laboratory for Neuro Imaging at the University of Southern California.

### Corticobasal Degeneration Study (sa000009) data

CBD Solutions funded the WES, data processing, and analysis. Assembled samples are from University College London (John Hardy), Mayo Clinic Jacksonville (Dennis Dickson), University of Pennsylvania (John Trojanowski), Emory University (Marla Gearing), Johns Hopkins University (Alex Pantelyat), Indiana University (Bernadino Ghetti), New York Brain Bank (Jean Paul Vonsattel), McClean Brain Bank (Elaine Benes), University of Texas Southwestern (Charles White), University of California Los Angeles (William Tourtelloute), and European collaborators at University Munich and Neurobiobank Munich (Gunter Hoglinger, Ulrich Muller, Hans Kretzschmr), Newcastle University, University of Barcelona (Charles Gaig), MRC London Brain Bank, Australian Brain Bank, and the University of Madrid (Alberto Rábano Gutiérrez).

### Progressive Supranuclear Palsy Study (sa000010) data

This work was funded by the following NIH grants: P01 AG017586 (VM-YL, GDS, JQT), U54 NS100693 (OR, DD, GDS), UG3 NS104095 (GDS, L-SW, OR), U54 AG052427 (l-SW, GDS), P30 AG010133 (B.G.), R01 AG057516 (AC, AM, AW, JAP, SG), R01 HL143790 (AC, SG), R01 HG010067 (SG), RF1 AG055477 (CB), P01 AG017586 (VM-YL, GDS, JQT, VMV), UG3 NS104095 and CWOW grant U54 NS100693 (DD), AG025688 and NS055077 (MG), P30 AG012300 (CLW), P30 AG053760 (APL and RA), 1P50NS091856 (RA), 5 P50 AG005134 (MPF), AG005131 (DRG), Johns Hopkins University Morris K. Udall Parkinson’s Disease Research Center of Excellence grant P50 NS038377 and Alzheimer’s Disease Research Center grant P50 AG05146 (JCT), U24 NS072026 and P30 AG19610 (TGB). This work was also funded by Cure PSP (GDS), the Rainwater Foundation (GDS), the Daniel B. Burke Endowed Chair for Diabetes Research (SG), the CHOP Center for Spatial and Functional Genomics (AW, SFG), a CUREPSP research grant (Cure PSP Grant # 515-14; 2013-2015) to P.P., the Reta Lila Weston Trust for Medical Research, the PSP Association (RdS), and the Michael J. Fox Foundation for Parkinson’s Research (TGB). G. Höglinger was funded by the German Research Foundation (DFG) under Germany’s Excellence Strategy within the framework of the Munich Cluster for Systems Neurology (EXC 2145 SyNergy – ID 390857198), the German Federal Ministry of Education and Research (BMBF, 01KU1403A EpiPD; 01EK1605A HitTau), and the NOMIS foundation (FTLD project). J. Hardy was partly funded by UKDRI limited which receives its funding from the MRC, the Alzheimer’s Society and Alzheimer Research UK. The London Neurodegenerative Diseases Brain Bank receives funding from the UK Medical Research Council (MR/L016397/1) and as part of the Brains for Dementia Research programme, jointly funded by Alzheimer’s Research UK and the Alzheimer’s Society. Queen Square Brain Bank is supported by the Reta Lila Weston Institute for Neurological Studies and the Medical Research Council UK. Newcastle Brain Tissue Resource is funded in part by a grant from the UK Medical Research Council (MR/L016451/1) and by Brains for Dementia Research, a joint venture between Alzheimer’s Society and Alzheimer’s Research UK (CMM) and National Institute of Health Research Biomedical Research Centre at Newcastle upon Tyne Hospitals NHS Foundation Trust and Newcastle University (CMM). This work was partly funded by UKDRI limited which receives its funding from the MRC, the Alzheimer’s Society and Alzheimer Research UK (JH). The Mayo Clinic Florida had support from a Morris K. Udall Parkinson’s Disease Research Center of Excellence (NINDS P50 #NS072187), CurePSP and the Tau Consortium. OAR is supported by a NINDS Tau Center without Walls (U54-NS100693), NINDS R01-NS078086 and the Mayo Clinic Center for Individualized Medicine. Funding provided by CurePSP through the generous support of the Peebler PSP Research Foundation in memory of Charles D. Peebler Jr. and Drs. Jeffrey S. and Jennifer R. Friedman in memory of Morton L. Friedman.

### Accelerating Medicines Partnership-Alzheimer’s Disease (AMP-AD) (sa000011) data

#### Mayo RNAseq Study

Study data were provided by the following sources: The Mayo Clinic Alzheimer’s Disease Genetic Studies, led by Dr. Nilufer Ertekin-Taner and Dr. Steven G. Younkin, Mayo Clinic, Jacksonville, FL using samples from the Mayo Clinic Study of Aging, the Mayo Clinic Alzheimer’s Disease Research Center, and the Mayo Clinic Brain Bank. Data collection was supported through funding by NIA grants P50 AG016574, R01 AG032990, U01 AG046139, R01 AG018023, U01 AG006576, U01 AG006786, R01 AG025711, R01 AG017216, R01 AG003949, NINDS grant R01 NS080820, CurePSP Foundation, and support from Mayo Foundation. Study data includes samples collected through the Sun Health Research Institute Brain and Body Donation Program of Sun City, Arizona. The Brain and Body Donation Program is supported by the National Institute of Neurological Disorders and Stroke (U24 NS072026 National Brain and Tissue Resource for Parkinson’s Disease and Related Disorders), the National Institute on Aging (P30 AG19610 Arizona Alzheimer’s Disease Core Center), the Arizona Department of Health Services (contract 211002, Arizona Alzheimer’s Research Center), the Arizona Biomedical Research Commission (contracts 4001, 0011, 05-901 and 1001 to the Arizona Parkinson’s Disease Consortium) and the Michael J. Fox Foundation for Parkinson’s Research

#### ROSMAP

We are grateful to the participants in the Religious Order Study, the Memory and Aging Project. This work is supported by the US National Institutes of Health [U01 AG046152, R01 AG043617, R01 AG042210, R01 AG036042, R01 AG036836, R01 AG032990, R01 AG18023, RC2 AG036547, P50 AG016574, U01 ES017155, KL2 RR024151, K25 AG041906-01, R01 AG30146, P30 AG10161, R01 AG17917, R01 AG15819, K08 AG034290, P30 AG10161 and R01 AG11101.

#### Mount Sinai Brain Bank (MSBB)

This work was supported by the grants R01AG046170, RF1AG054014, RF1AG057440 and R01AG057907 from the NIH/National Institute on Aging (NIA). R01AG046170 is a component of the AMP-AD Target Discovery and Preclinical Validation Project. Brain tissue collection and characterization was supported by NIH HHSN271201300031C.

### University of Pittsburgh- Kamboh (sa000012) data

This study was supported by the National Institute on Aging (NIA) grants AG030653, AG041718, AG064877 and P30-AG066468.

### APOE Extremes WGS Study (sa000013) data

We would like to thank study participants, their families, and the sample collectors for their invaluable contributions. This research was supported in part by the National Institute on Aging grant U01AG049508 (PI Alison M. Goate) and U01AG052411 (MPIs Alison Goate, Carlos Cruchaga, Bin Zhang). This research was supported in part by the National Institute on Aging Intramural Program (MPIs Sonja Scholz, Bryan Traynor, Andrew Singleton). This research was supported in part by Genentech, Inc. (PI Alison Goate, Robert Graham).

The NACC database is funded by NIA/NIH Grant U01 AG016976. NACC data are contributed by these NIA-funded ADCs: P30 AG013846 (PI Neil Kowall, MD), P50 AG008702 (PI Scott Small, MD), P50 AG025688 (PI Allan Levey, MD, PhD), P30 AG010133 (PI Andrew Saykin, PsyD), P50 AG005146 (PI Marilyn Albert, PhD), P50 AG005134 (PI Bradley Hyman, MD, PhD), P50 AG016574 (PI Ronald Petersen, MD, PhD), P30 AG013854 (PI M. Marsel Mesulam, MD), P30 AG008017 (PI Jeffrey Kaye, MD), P30 AG010161 (PI David Bennett, MD), P30 AG010129 (PI Charles DeCarli, MD), P50 AG016573 (PI Frank LaFerla, PhD), P50 AG005131 (PI Douglas Galasko, MD), P30 AG028383 (PI Linda Van Eldik, PhD), P30 AG010124 (PI John Trojanowski, MD, PhD), P50 AG005142 (PI Helena Chui, MD), P30 AG012300 (PI Roger Rosenberg, MD), P50 AG005136 (PI Thomas Grabowski, MD), P50 AG005681 (PI John Morris, MD), P30 AG028377 (Kathleen Welsh-Bohmer, PhD), and P50 AG008671 (PI Henry Paulson, MD, PhD). Samples from the National Cell Repository for Alzheimer’s Disease (NCRAD), which receives government support under a cooperative agreement grant (U24 AG21886) awarded by the National Institute on Aging (NIA), were used in this study. We thank contributors who collected samples used in this study, as well as patients and their families, whose help and participation made this work possible.

The Alzheimer’s Disease Genetics Consortium supported the collection of samples used in this study through National Institute on Aging (NIA) grants U01AG032984 and RC2AG036528.

### Cache County Study (sa000014) data

We acknowledge the generous contributions of the Cache County Memory Study participants. Sequencing for this study was funded by RF1AG054052 (PI: John S.K. Kauwe).

### NIH, CurePSP and Tau Consortium PSP WGS (sa000015) data

This project was funded by the NIH grant UG3NS104095 and supported by grants U54NS100693 and U54AG052427. Queen Square Brain Bank is supported by the Reta Lila Weston Institute for Neurological Studies and the Medical Research Council UK. The Mayo Clinic Florida had support from a Morris K. Udall Parkinson’s Disease Research Center of Excellence (NINDS P50 #NS072187), CurePSP and the Tau Consortium. The samples from the University of Pennsylvania are supported by NIA grant P01AG017586.

### CurePSP and Tau Consortium PSP WGS (sa000016) data

This project was funded by the Tau Consortium, Rainwater Charitable Foundation, and CurePSP. It was also supported by NINDS grant U54NS100693 and NIA grants U54NS100693 and U54AG052427. Queen Square Brain Bank is supported by the Reta Lila Weston Institute for Neurological Studies and the Medical Research Council UK. The Mayo Clinic Florida had support from a Morris K. Udall Parkinson’s Disease Research Center of Excellence (NINDS P50 #NS072187), CurePSP and the Tau Consortium. The samples from the University of Pennsylvania are supported by NIA grant P01AG017586. Tissues were received from the Victorian Brain Bank, supported by The Florey Institute of Neuroscience and Mental Health, The Alfred and the Victorian Forensic Institute of Medicine and funded in part by Parkinson’s Victoria and MND Victoria. We are grateful to the Sun Health Research Institute Brain and Body Donation Program of Sun City, Arizona for the provision of human biological materials (or specific description, e.g. brain tissue, cerebrospinal fluid). The Brain and Body Donation Program is supported by the National Institute of Neurological Disorders and Stroke (U24 NS072026 National Brain and Tissue Resource for Parkinson’s Disease and Related Disorders), the National Institute on Aging (P30 AG19610 Arizona Alzheimer’s Disease Core Center), the Arizona Department of Health Services (contract 211002, Arizona Alzheimer’s Research Center), the Arizona Biomedical Research Commission (contracts 4001, 0011, 05-901 and 1001 to the Arizona Parkinson’s Disease Consortium) and the Michael J. Fox Foundation for Parkinson’s Research. Biomaterial was provided by the Study Group DESCRIBE of theClinical Research of the German Center for Neurodegenerative Diseases (DZNE).

### UCLA Progressive Supranuclear Palsy (sa000017) data

We thank the AL-108-231 investigators for their contribution to this consortium dataset.

### Diagnostic Assessment of Dementia for the Longitudinal Aging Study of India (LASI-DAD) (sa000019) data

The Longitudinal Aging Study in India, Diagnostic Assessment of Dementia data is sponsored by the National Institute on Aging (grant numbers R01AG051125 and U01AG064948) and is conducted by the University of Southern California.

### Dissecting the Genomic Etiology of non-Mendelian Early-Onset Alzheimer Disease (EOAD) and Related Phenotypes (sa000023) data

This work was supported by the National Institutes of Health (NIH) grant R01AG064614. The ADSP-FUS is supported by U01AG057659.

The National Institutes of Health, National Institute on Aging (NIH-NIA) supported this work through the following grants: ADGC, U01 AG032984, RC2 AG036528; samples from the National Centralized Repository for Alzheimer’s Disease and Related Dementias (NCRAD), which receives government support under a cooperative agreement grant (U24 AG21886) awarded by the National Institute on Aging (NIA), were used in this study. Sequencing data generation and harmonization is supported by the Genome Center for Alzheimer’s Disease, U54AG052427, and data sharing is supported by NIAGADS, U24AG041689. We thank contributors who collected samples used in this study, as well as patients and their families, whose help and participation made this work possible.

NIH grants supported enrollment and data collection for the individual studies including the Alzheimer’s Disease Centers (ADC, P30 AG062429 (PI James Brewer, MD, PhD), P30 AG066468 (PI Oscar Lopez, MD), P30 AG062421 (PI Bradley Hyman, MD, PhD), P30 AG066509 (PI Thomas Grabowski, MD), P30 AG066514 (PI Mary Sano, PhD), P30 AG066530 (PI Helena Chui, MD), P30 AG066507 (PI Marilyn Albert, PhD), P30 AG066444 (PI John Morris, MD), P30 AG066518 (PI Jeffrey Kaye, MD), P30 AG066512 (PI Thomas Wisniewski, MD), P30 AG066462 (PI Scott Small, MD), P30 AG072979 (PI David Wolk, MD), P30 AG072972 (PI Charles DeCarli, MD), P30 AG072976 (PI Andrew Saykin, PsyD), P30 AG072975 (PI David Bennett, MD), P30 AG072978 (PI Neil Kowall, MD), P30 AG072977 (PI Robert Vassar, PhD), P30 AG066519 (PI Frank LaFerla, PhD), P30 AG062677 (PI Ronald Petersen, MD, PhD), P30 AG079280 (PI Eric Reiman, MD), P30 AG062422 (PI Gil Rabinovici, MD), P30 AG066511 (PI Allan Levey, MD, PhD), P30 AG072946 (PI Linda Van Eldik, PhD), P30 AG062715 (PI Sanjay Asthana, MD, FRCP), P30 AG072973 (PI Russell Swerdlow, MD), P30 AG066506 (PI Todd Golde, MD, PhD), P30 AG066508 (PI Stephen Strittmatter, MD, PhD), P30 AG066515 (PI Victor Henderson, MD, MS), P30 AG072947 (PI Suzanne Craft, PhD), P30 AG072931 (PI Henry Paulson, MD, PhD), P30 AG066546 (PI Sudha Seshadri, MD), P20 AG068024 (PI Erik Roberson, MD, PhD), P20 AG068053 (PI Justin Miller, PhD), P20 AG068077 (PI Gary Rosenberg, MD), P20 AG068082 (PI Angela Jefferson, PhD), P30 AG072958 (PI Heather Whitson, MD), P30 AG072959 (PI James Leverenz, MD). The Miami ascertainment and research were supported in part through: RF1AG054080, R01AG027944, R01AG019085, R01AG028786-02, RC2AG036528. The Columbia ascertainment and research were supported in part through: R37AG015473 and U24AG056270. The University of Washington ascertainment and research were supported in part through R01AG044546, RF1AG053303, RF1AG058501, U01AG058922 and R01AG064877.

### University of Alabama at Birmingham Alzheimer’s Disease Research Center (sa000036) data

Funding for genome sequencing was provided by the HudsonAlpha Memory and Mobility program. Funding for exome sequencing was provided by the Alzheimer’s Association. Funding for array genotyping was provided by NIA grant 5P20AG068024.

### Genetic Studies of Alzheimer’s Disease in Korea (sa000056) data

This research was supported by the Original Technology[HW1] [LF2] Research Program for Brain Science of the National Research Foundation (NRF) funded by the Korean government, MSIT (NRF-2014M3C7A1046041 and NRF-2014M3C7A1046042); by KBRI basic research program through Korea Brain Research Institute funded by the Ministry of Science and ICT (23-BR-03-05); by Healthcare AI Convergence Research & Development Program through the National IT Industry Promotion Agency of Korea (NIPA) funded by the Ministry of Science and ICT (No.1711120216); by National Institute on Aging grant U01-AG062602.

### Wellderly – Healthy Elderly Active Longevity (HEAL) (sa000057) data

Development of the Wellderly resource was supported by the National Center For Advancing Translational Sciences of the National Institutes of Health under Award Number UM1TR004407.

### Arizona APOE Cohort Study (sa000058) data

The Arizona APOE Cohort Study is conducted by Banner Alzheimer’s Institute located in Phoenix, AZ, USA in collaboration with Mayo Clinic Arizona in Scottsdale, AZ, USA. The Arizona APOE Cohort Study is supported by funding from the National Institute on Aging (NIA) of the National Institutes of Health (NIH) (R01AG069453). Research reported in this publication was also supported by the NIA under Award Number P30AG072980 to the Arizona Alzheimer’s Disease Research Center. We thank the staff and investigators of the studies as well as the participants whose help and participation made this work possible. The contents of this paper are solely the responsibility of the authors and do not necessarily represent the official views of the funders or study team.

### Genetic Architecture of Alzheimer’s disease and Related Proteinopathies (sa000059) data

This work was made possible by the support from the National Institutes of Health grants:

#### Pitt

AG064877, AG030653, AG041718, P30-AG066468, U01 AT000162, AG023651, AG052521, AG025516, UF1 AG051197, P01 AG025204, RF1 AG052525, AG052446.

#### WashU

R01AG044546, P01AG003991, RF1AG053303, R01AG058501, U01AG058922, RF1AG058501 and R01AG057777). The recruitment and clinical characterization of research participants at Washington University were supported by NIH P50 AG05681, P01 AG03991, and P01 AG026276. This work was supported by access to equipment made possible by the Hope Center for Neurological Disorders, and the Departments of Neurology and Psychiatry at Washington University School of Medicine.

We thank the contributors who collected samples used in this study, as well as patients and their families, whose help and participation made this work possible. This work was supported by access to equipment made possible by the Hope Center for Neurological Disorders, and the Departments of Neurology and Psychiatry at Washington University School of Medicine.

### ASPirin in Reducing Events in the Elderly (sa000060) data

ASPREE was supported by U01AG029824 from the National Institute on Aging (NIA) and the National Cancer Institute (NCI) at the National Institutes of Health (NIH), as well as grants 334047 and 1127060 from the National Health and Medical Research Council of Australia (NHMRC), and additional funding from Monash University and the Victorian Cancer Agency. ASPREE-XT is supported by U19AG062682 from NIA and NCI.

We thank the ASPREE/ASPREE-XT trial staff in Australia and the United States, the participants who volunteered for this trial, and the general practitioners and staff of the medical clinics who cared for the participants.

### Health & Aging Brain Study – Health Disparities (HABS-HD) (sa000061) data

Research reported in this publication was supported by the National Institute on Aging of the National Institutes of Health under Award Numbers R01AG054073 and R01AG058533, R01AG070862, P41EB015922 and U19AG078109. The content is solely the responsibility of the authors and does not necessarily represent the official views of the National Institutes of Health. We gratefully acknowledge the contributions of our study partners and their families, whose help and participation made this work possible.

### Estudio Familiar de la Influencia Genetica en Alzheimer (EFIGA) (sa000062) data

Data collection for this project was supported by the Genetic Studies of Alzheimer’s disease in Caribbean Hispanics (EFIGA) funded by the National Institute on Aging (NIA) and by the National Institutes of Health (NIH) (5R37AG015473, RF1AG015473, R56AG051876, R01AG067501). We acknowledge the EFIGA study participants and the EFIGA research and support staff for their contributions to this study.

### National Institute of Aging Alzheimer’s Disease Family Based Study (NIA AD-FBS) (sa000063) data

The NIA-AD FBS study supported the collection of samples used in this study through National Institute on Aging (NIA) grants U24AG056270,U24AG026395, R01AG041797 and U24AG056270. We thank contributors, including the Alzheimer’s disease Centers who collected samples used in this study, as well as patients and their families, whose help and participation made this work possible.

## Author contributions

The following authors designed the study: N.D.B., D.M.J.. The following authors performed all analyses and wrote the manuscript: N.D.B., D.M.J. and E.K.. The following authors contributed to data processing and analyses: N.D.B., D.M.J., E.K., R.C.T., A.N.L., A.B.G., D.J., T.P., A.M.G, M.K., A.E.R.. The following authors contributed to quality control: N.D.B., D.M.J., E.K., R.C.T., M.K., T.P., A.E.R.. The following authors contributed to writing specific parts of the text and preparing figures and tables for this manuscript: N.D.B., D.M.J., E.K., R.C.T., T.P., A.N.L., A.B.G., D.J., N.G., M.K.. The following authors provided critical feedback to method development: N.D.B., D.M.J., E.K., R.C.T., and M.K.. .The following authors provided critical feedback to analyses results: N.D.B., D.M.J., E.K., R.C.T., N.G., M.K., A.E.R.. The following authors provided critical feedback to the writing of the manuscript: N.D.B., D.M.J., E.K., R.C.T., A.N.L., A.B.G., D.J., T.P., A.M.G., B.S.G., N.G., M.K., A.E.R..

## Declaration of generative AI and AI-assisted technologies in the writing process

During the preparation of this work, the authors used Claude and ChatGPT in order to ensure that there are no grammatical errors in the manuscript. After using this tool/service, the authors reviewed and edited the content as needed and take full responsibility for the content of the publication.

